# Persistence and detection of anti-SARS-CoV-2 antibodies: immunoassay heterogeneity and implications for serosurveillance

**DOI:** 10.1101/2021.03.16.21253710

**Authors:** Javier Perez-Saez, María-Eugenia Zaballa, Sabine Yerly, Diego O. Andrey, Benjamin Meyer, Isabella Eckerle, Jean-François Balavoine, François Chappuis, Didier Pittet, Didier Trono, Omar Kherad, Nicolas Vuilleumier, Laurent Kaiser, Idris Guessous, Silvia Stringhini, Andrew S Azman, for the Specchio-COVID19 Study Group

## Abstract

Serologic studies have been critical in tracking the evolution of the COVID-19 pandemic. The reliability of serologic studies for quantifying the proportion of the population that have been infected depends on the extent of antibody decay as well as on assay performance in detecting both recent and older infections. Data on anti-SARS-CoV-2 antibodies persistence remain sparse, especially from infected individuals with few to no symptoms. In a cohort of mostly mild/asymptomatic SARS-CoV-2-infected individuals tested with three widely-used immunoassays, antibodies persisted for at least 8 months after infection, although detection depended on immunoassay choice, with one of them missing up to 40% of past infections. Simulations reveal that without appropriate adjustment for time-varying assay sensitivity, seroprevalence surveys may underestimate infection rates. As the immune landscape becomes more complex with naturally-infected and vaccinated individuals, assay choice and appropriate assay-performance-adjustment will become even more important for the interpretation of serologic studies.

## Introduction

Serosurveys have played an important role during the COVID-19 pandemic by helping track the true extent of transmission in different populations (*1*–*4*), and estimating key epidemiologic indicators such as the infection fatality ratio (IFR) (*5*–*7*). More than 400 serosurveys were published by the end of 2020, using dozens of different immunoassays, designed to detect antibodies targeting primarily all or part of the spike (S) or nucleocapsid (N) proteins of the SARS-CoV-2 virus (*8*). The accuracy of serology-based estimates depends on the immunoassay antibody targets and their performance in detecting both recent and historic infections. At this stage of the pandemic, successive epidemic waves in different parts of the world create a diverse mix of people infected over different times in the past, challenging SARS-CoV-2 serosurveillance because of this increasingly heterogeneous immunological landscape.

Anti-SARS-CoV-2 antibody levels tend to decay after the convalescent period, which can lead to increasing chances that immunoassays provide a negative result (*9, 10*). When immunoassays are used as a proxy for historic infections, as typically done in serosurveys, these are considered false negatives. Furthermore, post-infection antibody kinetics appear to be differential by infection severity, with severe infections leading to larger increases in antibodies than mild or asymptomatic infections (*11*). False negatives can lead to severe underestimates of the true infection attack rate unless appropriately understood and accounted for in analyses (*12*). However, few studies have characterized antibody kinetics past six months after infection (*13, 14*), and few have described these kinetics in mild and asymptomatic infections (*15*–*18*), which comprise the vast majority of infections in the community (*19*). Quantifying the changes in antibody detection for available immunoassays across the spectrum of infection severity and age is therefore essential in providing robust estimates of key epidemiological features of the COVID-19 pandemic (*12, 20*).

Here we quantify changes in anti-SARS-CoV-2 antibody levels up to 9 months from plausible dates of infection in a cohort of seropositive individuals recruited through serosurveys conducted in Geneva, Switzerland. We use three different immunoassays to compare test performance, including the reduction in sensitivity as time since infection increases. Our results provide critical insights into the interpretation of population-based serologic studies as the immune landscape becomes more complex due to a mix of recent and distal infections and vaccination.

## Results

### Cohort recruitment and characteristics

For this study, participants recruited between April and July 2020 through serosurveys conducted in the canton of Geneva, Switzerland (*2, 29*), returned for a follow-up blood draw in November 2020 (Figure 1a). A total of 354 participants from those surveys that had a positive Euroimmun anti-S1 IgG (hereafter EI) test result at baseline constituted the EI-positive cohort. Participants in this cohort were aged between 18 and 84 years, and 52% (183/354) were women (Table 1). Less than half reported having had an RT-PCR test prior to the baseline visit (148, 42%), 58 of whom reported a positive result (90 negative, positivity rate of 39%). Ten percent (37/354) of participants reported having had no COVID-19-compatible symptoms before the baseline visit, while 69% reported 4 symptoms or more. The four most frequently reported COVID-19-compatible symptoms were fever (66%), fatigue (63%), headache (58%) and taste and smell loss (55%) (Figure S1). The majority of these participants did not require hospitalization (334/354, 94%), and only 2 required a stay in an ICU (0.6%, with missing information for 5 hospitalized participants). The median period between baseline and follow-up visits was 165 days (range: 115-224 days) (Figure 1b). Twenty percent (71/354) of participants reported having performed a SARS-CoV-2 virologic test (RT-PCR or rapid antigen) between visits including 4 (6%) reporting a positive result. No participant reported to have been hospitalized between their baseline and follow-up visits.

**Table 1.**
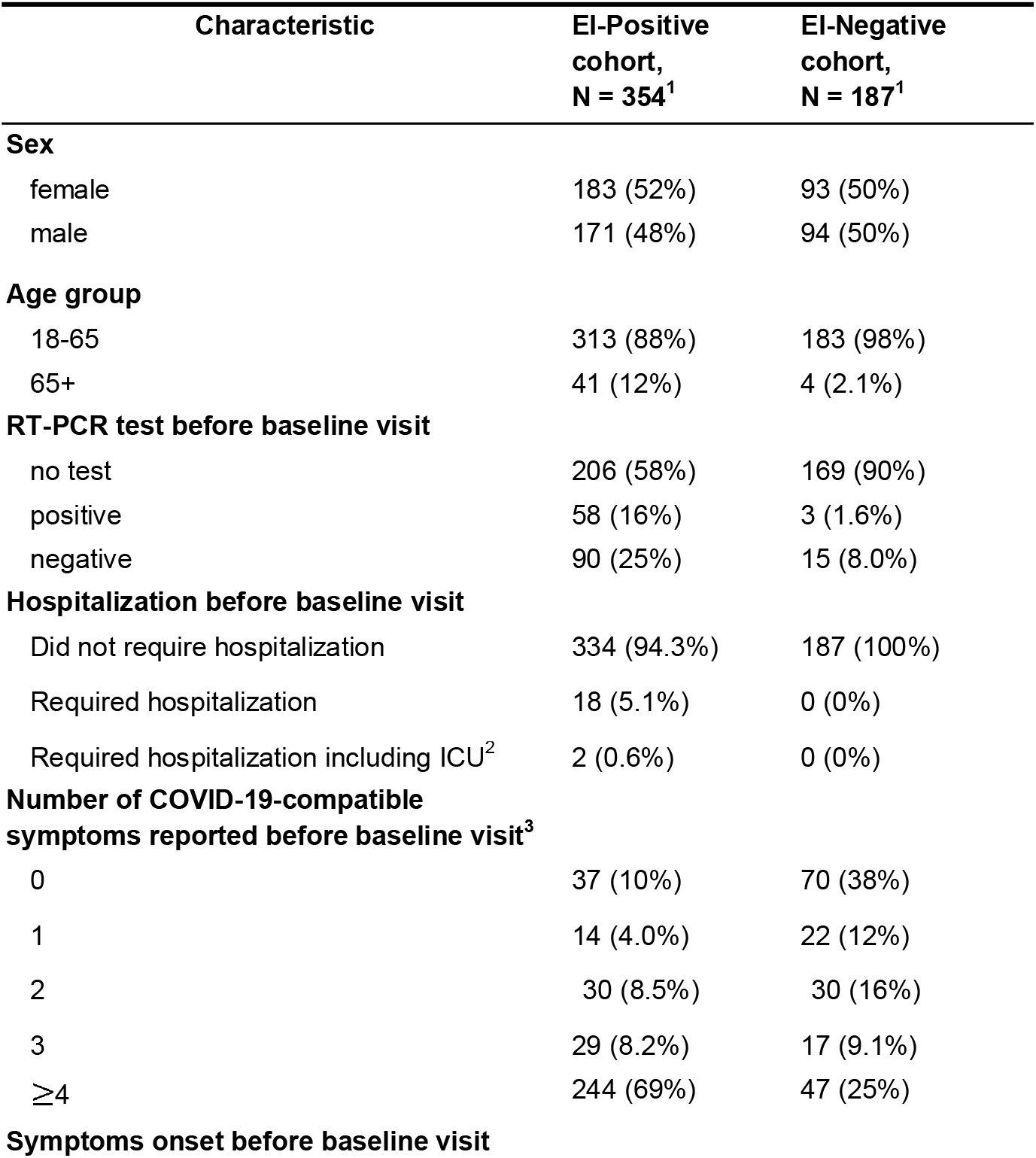
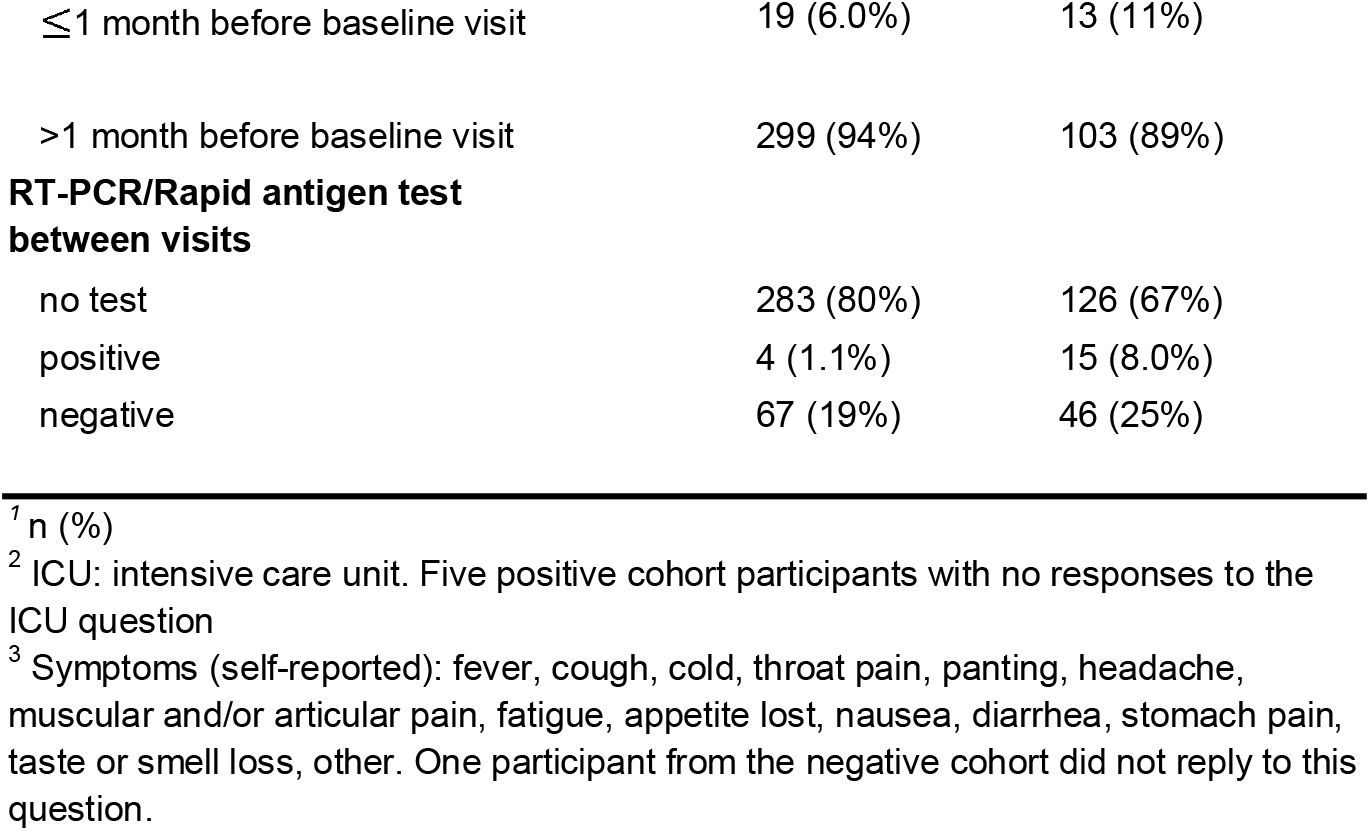
Characteristics of baseline Euroimmun anti-S1 IgG (EI) positive and negative cohorts.

**Figure 1:**
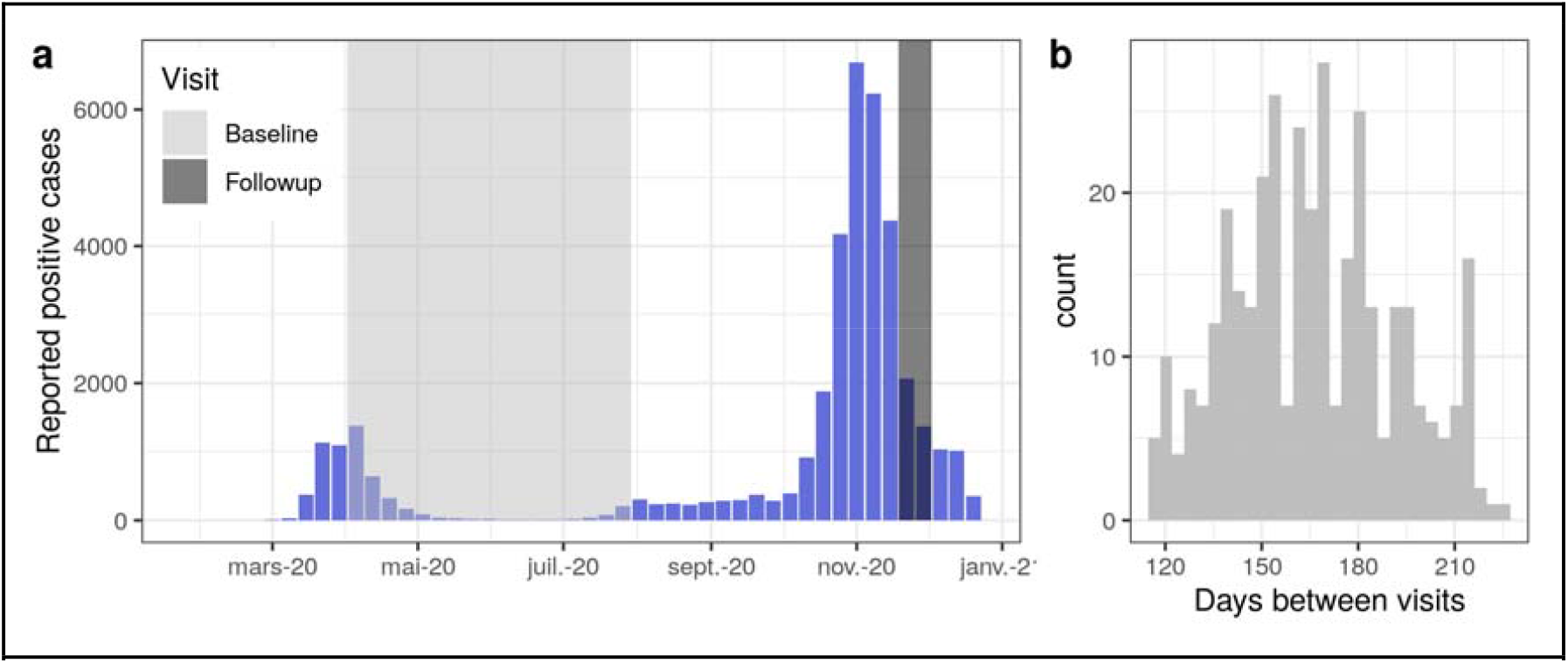
Study recruitment with respect to the SARS-CoV-2 epidemic curve in Geneva, Switzerland. a) Weekly reported number of virologically-confirmed SARS-CoV-2 infections in the canton of Geneva (blue bars) and study timing for both the baseline (light gray) and follow-up (dark gray) visits. b) Histogram of days between study visits for the EI-positive cohort (N=354).

**Figure 2:**
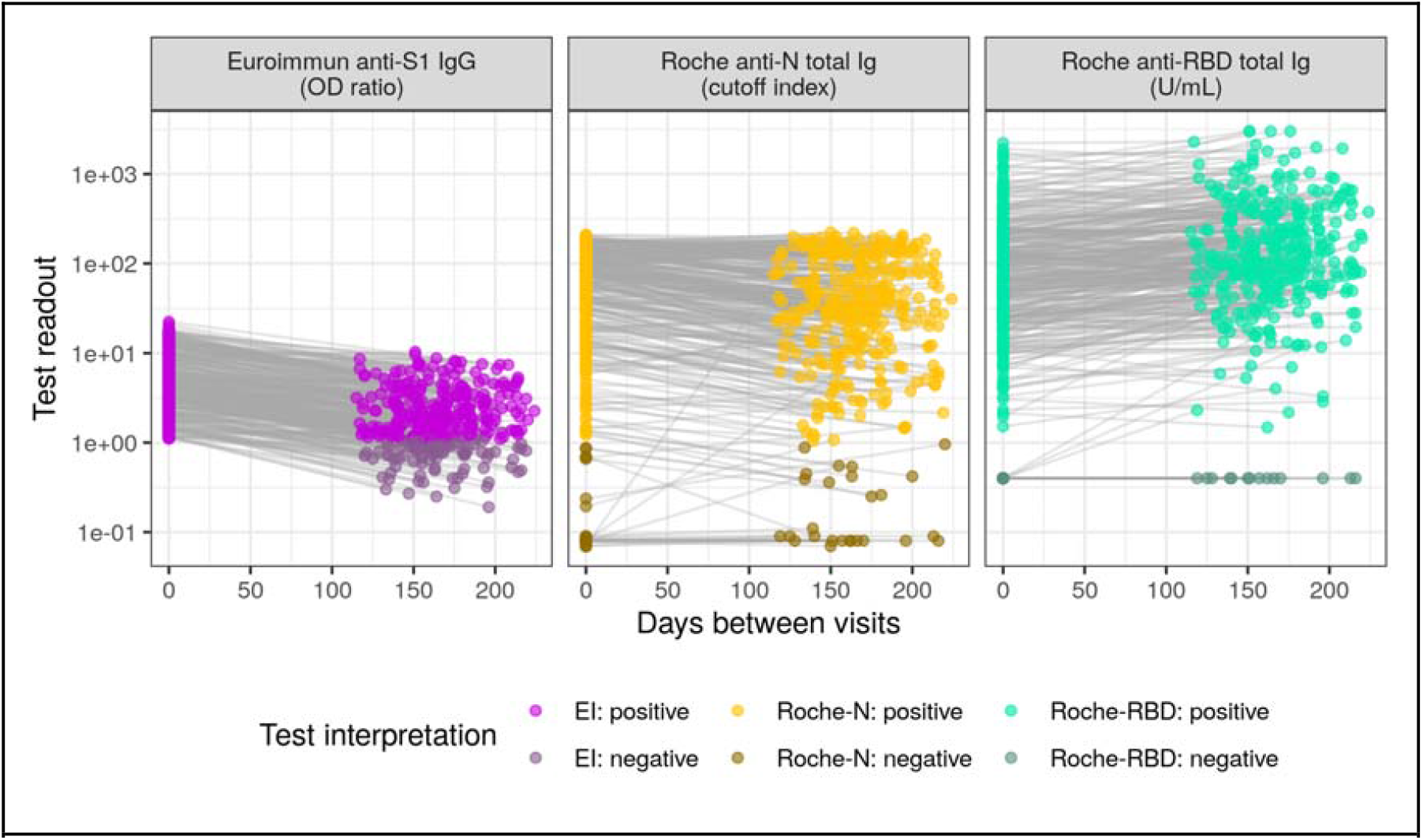
Test readout trajectories between baseline and follow-up visits. The cohort was composed of 354 participants with positive Euroimmun anti-S1 (EI) test at baseline.Test readout units and thresholds for positivity are assay-specific (Materials and Methods), Roche-RBD values below the limit of quantitation (0.4 U/mL) were set to the limit of quantitation for plotting and analysis. The dynamic range of both the EI and Roche-N tests are limited compared to the Roche-RBD thus leading to censoring of extremely high and low values. Baseline and follow-up samples were tested with different reagent lots of the EI immunoassay whereas the same Roche-N and Roche-RBD reagent lots were used for all samples (Appendix, section S2). Trajectories for the EI-negative cohort are given in Figure S3.

We also followed a cohort of 187 participants who had a negative EI test result at baseline (EI-negative cohort) selected from previous participants to have a similar sex ratio and age range as the EI-positive cohort (Table 1). Three individuals (3/18) in this EI-negative cohort reported having a positive RT-PCR test prior to the baseline visit. Thirty eight percent of participants reported having had no COVID-19-compatible symptoms and none reported having been hospitalized prior to the baseline visit. A total of 61 participants in the EI-negative cohort (33%) reported having had a SARS-CoV-2 virologic test (RT-PCR or rapid antigen) between visits with 15 (25%) reporting a positive result.

### Antibody detection and decay with three immunoassays

In addition to the Euroimmun anti-S1 IgG test, samples from both study visits were tested with two Roche total Ig assays, one targeting anti-RBD and the other anti-N antibodies (Materials and Methods). Within the EI-positive cohort, 93.2% (330/354) and 95.2% (337/354) were positive for the Roche-N and Roche-RBD assays, respectively (2-by-2 confusion matrices given in Fig. S2). At the individual-level, 26% (91/354) of those in the EI-positive cohort became seronegative with the EI test at followup (i.e., sero-reverted, Table 2). Sero-reversions were much less frequent with the Roche-N assay (1.2%, 6/330) and none were detected with the Roche-RBD assay. We identified no significant differences in the proportion of participants seroreverting across age groups and sex for all three tests (Table S1).

**Table 2:**
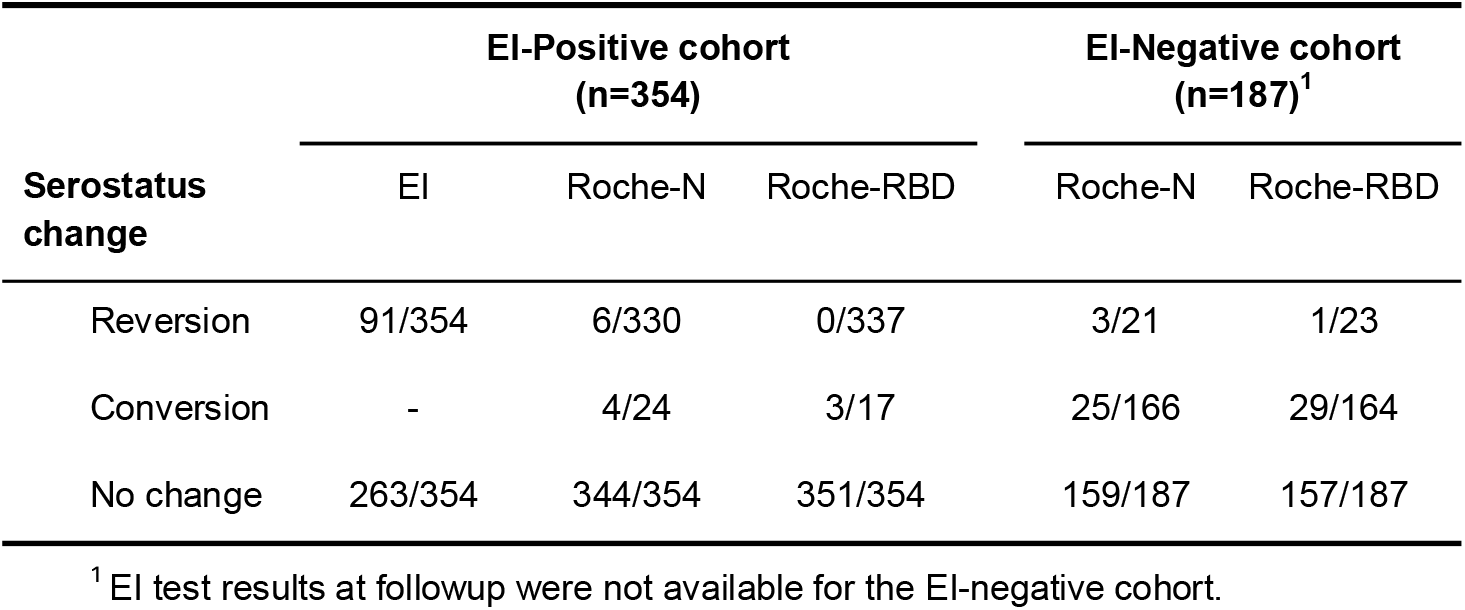
Serostatus and test readout changes between visits. Serostatus changes are given with respect to the baseline number of positives (negatives) for reversion (conversion) for each test. Statistics by sex and age group given in Table S1.

Beyond seroreversions, we quantified the change in test readout between visits for the Roche-RBD immunoassay, as the other two tests are considered qualitative or semiquantitative by the manufacturers. We found that 17% (61/354) of participants had a significant decrease in their test readout and 66% (235/354) had a significant increase. When subdivided by sex and age class, women had a significantly lower proportion of decaying as well as a higher proportion of increasing Roche-RBD responses, and no differences were found between age classes (Table S1).

In the EI-negative cohort, around 10% of the participants were classified as seropositive at baseline based on the Roche-N (21/187) and Roche-RBD (23/187) assays (Figure S2). A total of 25 out of 166 (15%) Roche-N negatives, and 29/164 (17.5%) Roche-RBD negatives became seropositive at follow-up (Table 2). We identified no significant differences by age or sex in seroconversion rates (Table S1).

### Estimation of time-varying test sensitivity and impact on serosurveillance

The trajectories of antibody detection in both cohorts, combined with data from assay validation studies (Table S2, Fig. S7) enabled us to produce model-based estimates of test clinical specificity and changes in clinical sensitivity with time post infection (Materials and Methods). We estimate the specificity of the EI immunoassay to be 95.3% (95% Credible Interval, CrI: 93.5-96.7), lower than that of both Roche tests, which were close to 100% (Roche-N: 99.8%, 95% CrI:99.4-100; Roche-RBD: 99.6%, 95% CrI:98.3-100).

We estimated large differences in sensitivity between tests, which depended on the delay between infection and the date of serologic assessment (Fig. 3a). After an initial rise in the first few weeks post infection, sensitivity peaked after 52 days (95% CrI: 39-64) for EI (peak value 95.8%, 95% CrI: 93.9-97.5), 81 days (95% CrI: 60-96) for Roche-N (peak value 99.1%, 95% CrI: 98.3-99.7), an 269 days (95% CrI: 122-284) for Roche-RBD (peak value 99.8%, 95% CrI: 99.3-100). EI sensitivity then decreased gradually reaching 61.2% (95% CrI: 53.4-68.5) at 284 days post infection, the longest time modeled. As baseline and follow-up samples were tested with different EI test reagent lots, we also estimated performance where we restricted to 127 samples with baseline and follow-up reagent lots with similar test readout values for our internal positive control serum and found similar results for sensitivity and the proportion who seroreverted, though with larger uncertainty (Figure 3a, Supplementary Appendix section S2). The decrease in sensitivity for Roche-N was smaller (mean at 284 days of 93.3%, 95% CrI: 88.7-96.7). Sensitivity estimates of Roche-RBD remained close to the peak value up to the maximal modeled time, with a 88% posterior probability that sensitivity was still increasing after 284 days. No significantly different results were obtained for both Roche tests when restricting the analysis to the 127 samples mentioned above (Fig. S8). We note that these estimates account for the 17.4% (95% CrI: 11.9-23.6) probability of SARS-CoV-2 infection between visits that is jointly estimated in the modelling framework (Materials and Methods, Appendix section S3).

**Figure 3:**
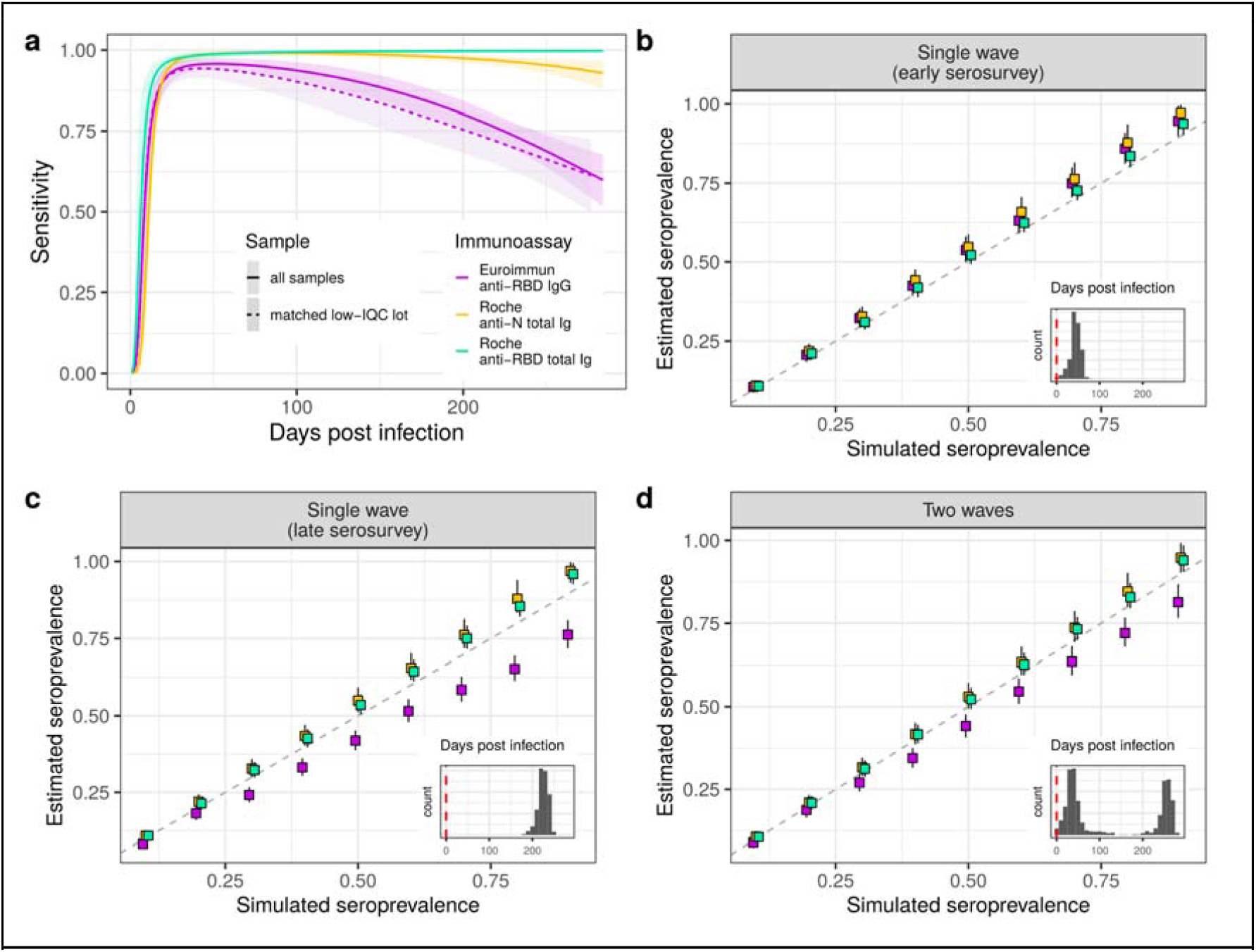
Model test performance estimates and simulation. a) Model estimates of sensitivity changes with time post infection. Due to EI reagent inter-lot variability (Supplementary Appendix Section S2) results are shown for the whole sample (N = 354), as well as for a subsample for which assay internal positive quality control (IQC) readout values were similar for baseline and follow-up reagent lots (N = 127, matched low IQC lot). b-c-d) Simulation scenarios of seroprevalence estimation if the decay in sensitivity is not accounted for. Scenario in b) is assumed to occur one month after the first epidemic wave peak in Geneva, with corresponding distribution of days between infection and the serosurvey ; scenario in c) the serosurvey occurs after a single wave and 180 days after the epidemic peak; and scenario in d) assumes the serosurvey occurred one month after the peak of the second epidemic wave, yielding a bimodal distribution of days post infection (insets, vertical dashed line at x=0 indicates infections that occurred on the serosurvey date).

To explore the potential implications of not accounting for time-varying sensitivity when estimating the proportion of the population infected through serosurveys, we simulated an array of serosurveys with different shapes and using different immunoassays. We simulated serosurveys in epidemics shaped like that of Geneva, and conducted either i) shortly after a single wave as the one in spring 2020, ii) long after this single wave, or iii) after two successive waves (Materials and Methods). When serosurveys were simulated just after a single epidemic wave (i), with times post-infection ranging from 0 to 115 days (Fig. 3b), seroprevalence estimates for all three tests using conventional adjustment for sensitivity have 95% credible intervals covering the true values of simulated seroprevalence between 10 and 30%, after which estimates had a slight tendency towards overestimation. Instead, when simulating serosurveys longer after a single wave (180-250 days post-infection), as the true underlying seroprevalence increased, estimates based on the EI assay grew increasingly biased, with an under-estimation of up to 15% when the simulated seroprevalence was 90% (Fig. 3c). In contrast, estimates based on the two Roche assays tended to overestimate seroprevalence by 5% in this scenario. Finally, the results obtained for the two epidemic waves scenario stood in between, with a less severe seroprevalence underestimation for EI test (10% underestimation at 90% seroprevalence). The two Roche tests remained in closer agreement with the true seroprevalence throughout the simulated range with the 95% CrI covering the true value at seroprevalence below 30%, and slightly overestimating by around 3% it for higher seroprevalence (>60%).

## Discussion

In a cohort of mostly mild/asymptomatic SARS-CoV-2-infected individuals, we found that antibodies targeting either the nucleocapsid (N) or the spike (S) proteins of the virus generally persist for at least 8 months after infection, although their detection depends on immunoassay choice. We found that the initial measurements taken within 4.5 months of participants’ infections were consistent across the three assays used. However, results diverged between assays upon re-evaluation 4-8 months later, with about one-in-four participants sero-reverting according to the EI IgG assay, as opposed to the Roche anti-N and anti-RBD total Ig tests, which respectively detected only few and no sero-reversions. Through simulation analyses, we show that without appropriate quantification and adjustment for time-varying assay sensitivity, seroprevalence surveys may underestimate the true number of cumulative infections in a population.

The persistence of seropositivity in this cohort of mostly mild infections provides encouraging prospects for the continued use and interpretation of population-based serosurveys to track the progress of the pandemic. However, we show that the differences in seropositivity rates between commercially-available tests that had been observed in the early phase of the COVID-19 pandemic can be amplified at longer follow-up times (*21*–*23*). Decaying assay sensitivity like the one we observed here for the EI assay may lead to the under-estimation of cumulative attack rates as shown by our simulations, and seroprevalence corrections may be warranted (*4*). We note that the simulations in this work are based on the epidemic curve in Geneva, Switzerland, and that seroprevalence estimation bias caused by decaying sensitivity will depend on the timing of serosurveillance with respect to the number and amplitude of preceding epidemic waves, with larger and more distal waves being more severely under-estimated. Even for mild infections, which are thought to elicit less robust immune responses (*24*), the sensitivity of anti-RBD and anti-N total Ig Roche tests remained close to 100% after more than 8 months post-infection. These results suggest that both Roche immunoassays are suitable for seroprevalence estimation at longer times post-exposure.

From an immunological perspective these results raise questions related to the dynamics and duration of immunity against SARS-CoV-2 infection. Previous studies suggest that anti-RBD antibody measurement correlates with neutralization titers at least up to 4 to 6 months post exposure (*11, 25*). Given that we did not perform neutralization assays, it remains unclear whether the persistence of antibodies we report here are a proxy of continued immune protection. We also highlight the discrepancy between the decrease of anti-S1 IgG as measured by the Euroimmun assay and the increase of total anti-RBD Ig measured by the Roche-RBD assay, the latter having been observed in other studies using the same test, which however correlated poorly with neutralizing antibody measurements (*16*). The increase in readout values from the Roche-RBD assay, and to a lesser extent for Roche-N, over time for many participants may be explained by aspects of the assay design which leads to preferential binding of higher avidity antibodies (which increase after infection) or due to the fact that they are measuring total immunoglobulins versus a single isotype (*26*).

Our results come with a number of limitations. While our estimates of seroreversion with the EI assay were in line with previously published data (*10, 27, 28*), we tested baseline and follow-up samples with the EI assay at different times and with different reagent lots (inter-lot coefficient of variation of 30%, Fig. S4). Furthermore, our internal quality control (pooled COVID-19 positive patient serum) in the follow-up lot had significantly lower readout values than most lots used for baseline samples (Supplementary Appendix section S2). To explore the potential effects of this lot-to-lot variability in our assessment of EI performance over time, we conducted sensitivity analyses by matching tests run in one baseline batch that had similarly low readout estimates as the follow-up batch (Fig. S5) and found similar estimates of the proportion who sero-reverted and time-varying sensitivity (Fig. 3a and Supplement). While this reagent inter-lot variability did not appear to have great impacts on our specific results, standardization of readout values (e.g., through the use of monoclonal antibodies) can help ensure compatibility between lots and labs. Secondly, statistics on changes in sero-status and test response may have been influenced by (re-)exposures during the period between baseline and follow-up visits. We attempted to account for the observed 17% seroconversion rate among initially seronegatives in the model of time-varying sensitivity but were unable to do so explicitly in other analyses. Finally the cohorts in this study did not include children, whose long-term antibody dynamics have not yet been well documented in the published literature.

Through quantifying anti-SARS-CoV-2 antibody persistence in a cohort of mostly mildly symptomatic and asymptomatic infections, we confirm that antibodies remain detectable after at least 8 months post-infection. Using multiple immunoassays, we illustrate that test choice matters and can greatly affect the interpretation of results from population-level serologic studies, especially as the immune landscape becomes a more complex mix of recent and old infections. While the Roche anti-RBD total Ig assay used in these analyses appears to have excellent performance across the spectrum of clinical severity over time, measurement of antibody responses targeting the spike RBD alone in partially vaccinated populations will be difficult to interpret and multi-epitope tests may become increasingly useful. Continued multi-assay, multi-epitope characterization of post-infection kinetics over longer periods can help continue to allow for appropriate analyses and interpretation of data from serosurveillance efforts aimed at tracking the evolution of this pandemic.

## Materials and Methods

### Recruitment

The results presented here correspond to the follow-up of participants recruited between April and July 2020 at one of two previous serosurveys: SEROCoV-POP (*2*) and SEROCoV-WORK+ (29). Selected participants from these two studies who were seropositive on the Euroimmun anti-S1 test (OD ratio ≥ 1.1, EI-positive cohort) at their first study visit (referred to as ‘baseline’) were invited to return for another serologic test in November 2020 (referred to as ‘follow-up’). To estimate the infection risk in the community over the period between the two study visits, we also selected 187 participants initially negative on the Euroimmun anti-S1 test with similar sex ratio and age range who returned for a second visit. Procedures in all cases (first two serosurveys for baseline visit and follow-up visit) were similar: all participants gave written informed consent, completed a questionnaire and provided a venous blood sample. This study was approved by the Geneva Cantonal Commission for Research Ethics (CCER project number 2020-00881).

### Immunoassays

SARS-CoV-2 antibodies were measured using three commercially-available tests: a semiquantitative anti-S1 ELISA detecting IgG (Euroimmun, Lübeck, Germany #EI 2606-9601 G, referred to as EI), and the quantitative Elecsys anti-RBD (#09 289 275 190, Roche-S) and semiquantitative Elecsys anti-N (#09 203 079 190, Roche-N), both measuring IgG/A/M levels (Roche Diagnostics, Rotkreuz, Switzerland). In each case, seropositivity was defined using the cut-off provided by the manufacturer: ratio ≥ 1.1 for Euroimmun anti-S1 ELISA; titer ≥ 0.8 U/mL for Roche anti-RBD; and cut-off index ≥ 1.0 for Roche anti-N. The Euroimmun immunoassay on samples from the baseline and follow-up visits was performed as samples came into the lab over the course of the studies, using several different reagent lots. On the other hand, all Roche tests (anti-RBD and anti-N) on baseline and follow-up samples were performed using the same reagent lot in each case. Samples from the baseline visit were frozen after their first analysis and thawed several months later for retesting with the Roche immunoassays (Supplementary Appendix section S2).

### Statistical Analyses

We determined the proportion who seroconverted (negative to positive) or seroreverted (positive to negative), and test the significance in proportions between sex and age class using a 2-sample test for equality of proportions with continuity correction (Table S1). We also compared test readout values at each visit and classified each participant’s response as decreasing, increasing or stable for Roche-RBD test readouts (the only quantitative test in this study). We assessed significance of changes in test readout values between visits taking into account the inter-lot variance of our internal positive control serum. The coefficient of variation of the Roche-RBD test readout was set to 7.6% (Supplementary Fig. S4). Significance of response changes was based on the z-score of the difference between follow-up and baseline results at a significance level of 5%.

We developed a statistical model to jointly infer each test’s specificity and sensitivity accounting both for changes in sensitivity with time post-infection due to antibody decay as well as possible unknown SARS-CoV-2 infection times including the possibility of infection between visits (Fig. S6). Inference is drawn in a Bayesian framework that allows to incorporate multiple sources of test validation data as well as RT-PCR test results when available (details given in Supplementary Appendix, sections S3 and S4).

We then used simulations to illustrate how seroprevalence estimates could be biased if only correcting for sensitivity and specificity using the Gland-Rogen estimator (30) with data from typical validation studies in the literature and package inserts (single time invariant sensitivity, short follow-up times and more representative of severe infections). We considered three hypothetical scenarios of serosurveillance sampling using the infection histories associated with Geneva’s epidemic curve as an example, the first one occurring one month after the peak of the first wave, the second one as if it had occurred five months after the single wave, and the third one month after the peak of the second wave (Fig. 1). This results in distinct distributions of time since infection, unimodal around short (∼1 month) and longer (∼5 months) times post infection for the first and second, and bimodal with a peak at short and one at longer times (∼8 months) post infection for the third. We then used these distributions to simulate test results based on our estimates of specificity and time-varying test sensitivity. We finally estimated the seroprevalence correcting for test performance, but using the conventional approach with a single value for sensitivity, inferred in a Bayesian hierarchical framework that incorporates multiple validation sources (31). For each scenario we simulated 2000 samples with seroprevalence ranging from 10 to 90% and compared these simulated data to the seroprevalence estimates that ignore changes in sensitivity. Details on seroprevalence estimation using the “conventional” approach are described in the Supplementary Appendix section S4.

## Data Availability

Code are available at https://github.com/UEP-HUG/serosuivi-public along with dummy data. Data can be made available upon request by contacting the corresponding author.

## Acknowledgments

We thank all the participants, without whom this study would not have been possible.

## References

1. M. Pollán, B. Pérez-Gómez, R. Pastor-Barriuso, J. Oteo, M. A. Hernán, M. Pérez-Olmeda, J. L. Sanmartín,A. Fernández-García, I. Cruz, N. Fernández de Larrea, M. Molina, F. Rodríguez-Cabrera, M. Martín, P. Merino-Amador, J. León Paniagua, J. F. Muñoz-Montalvo, F. Blanco, R. Yotti, ENE-COVID Study Group, Prevalence of SARS-CoV-2 in Spain (ENE-COVID): a nationwide, population-based seroepidemiological study. Lancet. 396, 535–544 (2020).

2. S. Stringhini, A. Wisniak, G. Piumatti, A. S. Azman, S. A. Lauer, H. Baysson, D. De Ridder, D. Petrovic, S. Schrempft, K. Marcus, S. Yerly, I. Arm Vernez, O. Keiser, S. Hurst, K. M. Posfay-Barbe, D. Trono, D. Pittet, L. Gétaz, F. Chappuis, I. Eckerle, N. Vuilleumier, B. Meyer, A. Flahault, L. Kaiser, I. Guessous, Seroprevalence of anti-SARS-CoV-2 IgG antibodies in Geneva, Switzerland (SEROCoV-POP): a population-based study. Lancet. 396, 313–319 (2020).

3. I. Eckerle, B. Meyer, SARS-CoV-2 seroprevalence in COVID-19 hotspots. Lancet. 396 (2020), pp. 514–515.

4. L. F. Buss, C. A. Prete Jr, C. M. M. Abrahim, A. Mendrone Jr, T. Salomon, C. de Almeida-Neto, R. F. O. França, M. C. Belotti, M. P. S. S. Carvalho, A. G. Costa, M. A. E. Crispim, S. C. Ferreira, N. A. Fraiji, S. Gurzenda, C. Whittaker, L. T. Kamaura, P. L. Takecian, P. da Silva Peixoto, M. K. Oikawa, A. S. Nishiya, V. Rocha, N. A. Salles, A. A. de Souza Santos, M. A. da Silva, B. Custer, K. V. Parag, M. Barral-Netto, M. U. G. Kraemer, R. H. M. Pereira, O. G. Pybus, M. P. Busch, M. C. Castro, C. Dye, V. H. Nascimento, N. R. Faria, E. C. Sabino, Three-quarters attack rate of SARS-CoV-2 in the Brazilian Amazon during a largely unmitigated epidemic. Science. 371, 288–292 (2021).

5. J. Perez-Saez, S. A. Lauer, L. Kaiser, S. Regard, E. Delaporte, I. Guessous, S. Stringhini, A. S. Azman, Serocov-POP Study Group, Serology-informed estimates of SARS-CoV-2 infection fatality risk in Geneva, Switzerland. Lancet Infect. Dis. (2020), doi:10.1016/S1473-3099(20)30584-3.

6. O. B. Pedersen, J. Nissen, K. M. Dinh, M. Schwinn, K. A. Kaspersen, J. K. Boldsen, M. Didriksen, J. Dowsett, E. Sørensen, L. W. Thørner, M. A. H. Larsen, B. Grum-Schwensen, S. Sækmose, I. W. Paulsen, N. L. S. Frisk, T. Brodersen, L. S. Vestergaard, K. Rostgaard, K. Mølbak, R. L. Skov, C. Erikstrup, H. Ullum, H. Hjalgrim, SARS-CoV-2 infection fatality rate among elderly retired Danish blood donors - A cross-sectional study. Clin. Infect. Dis. (2020), doi:10.1093/cid/ciaa1627.

7. A. T. Levin, W. P. Hanage, N. Owusu-Boaitey, K. B. Cochran, S. P. Walsh, G. Meyerowitz-Katz, Assessing the age specificity of infection fatality rates for COVID-19: systematic review, meta-analysis, and public policy implications. Eur. J. Epidemiol. 35, 1123–1138 (2020).

8. X. Chen, Z. Chen, A. S. Azman, X. Deng, R. Sun, Z. Zhao, N. Zheng, X. Chen, W. Lu, T. Zhuang, J. Yang, C. Viboud, M. Ajelli, D. T. Leung, H. Yu, Serological evidence of human infection with SARS-CoV-2: a systematic review and meta-analysis. The Lancet Global Health (2021), doi:10.1016/S2214-109X(21)00026-7.

9. S. C. A. Nielsen, F. Yang, R. A. Hoh, K. J. L. Jackson, K. Roeltgen, J.-Y. Lee, A. Rustagi, A. J. Rogers, A. E. Powell, P. S. Kim, T. T. Wang, B. Pinsky, C. A. Blish, S. D. Boyd, B cell clonal expansion and convergent antibody responses to SARS-CoV-2. Res Sq (2020), doi:10.21203/rs.3.rs-27220/v1.

10. D. F. Gudbjartsson, G. L. Norddahl, P. Melsted, K. Gunnarsdottir, H. Holm, E. Eythorsson, A. O. Arnthorsson, D. Helgason, K. Bjarnadottir, R. F. Ingvarsson, B. Thorsteinsdottir, S. Kristjansdottir, K. Birgisdottir, A. M. Kristinsdottir, M. I. Sigurdsson, G. A. Arnadottir, E. V. Ivarsdottir, M. Andresdottir, F. Jonsson, A. B. Agustsdottir, J. Berglund, B. Eiriksdottir, R. Fridriksdottir, E. E. Gardarsdottir, M. Gottfredsson, O. S. Gretarsdottir, S. Gudmundsdottir, K. R. Gudmundsson, T. R. Gunnarsdottir, A. Gylfason, A. Helgason, B. O. Jensson, A. Jonasdottir, H. Jonsson, T. Kristjansson, K. G. Kristinsson, D. N. Magnusdottir, O. T. Magnusson, L. B. Olafsdottir, S. Rognvaldsson, L. le Roux, G. Sigmundsdottir, A. Sigurdsson, G. Sveinbjornsson, K. E. Sveinsdottir, M. Sveinsdottir, E. A. Thorarensen, B. Thorbjornsson, M. Thordardottir, J. Saemundsdottir, S. H. Kristjansson, K. S. Josefsdottir, G. Masson, G. Georgsson, M. Kristjansson, A. Moller, R. Palsson, T. Gudnason, U. Thorsteinsdottir, I. Jonsdottir, P. Sulem, K. Stefansson, Humoral Immune Response to SARS-CoV-2 in Iceland. N. Engl. J. Med. 383, 1724–1734 (2020).

11. K. Röltgen, A. E. Powell, O. F. Wirz, B. A. Stevens, C. A. Hogan, J. Najeeb, M. Hunter, H. Wang, M. K. Sahoo, C. Huang, F. Yamamoto, M. Manohar, J. Manalac, A. R. Otrelo-Cardoso, T. D. Pham, A. Rustagi, A. J. Rogers, N. H. Shah, C. A. Blish, J. R. Cochran, T. S. Jardetzky, J. L. Zehnder, T. T. Wang, B. Narasimhan, S. Gombar, R. Tibshirani, K. C. Nadeau, P. S. Kim, B. A. Pinsky, S. D. Boyd, Defining the features and duration of antibody responses to SARS-CoV-2 infection associated with disease severity and outcome. Sci Immunol. 5 (2020), doi:10.1126/sciimmunol.abe0240.

12. S. Takahashi, B. Greenhouse, I. Rodríguez-Barraquer, Are Seroprevalence Estimates for Severe Acute Respiratory Syndrome Coronavirus 2 Biased? J. Infect. Dis. 222, 1772–1775 (2020).

13. E. H. Y. Lau, O. T. Y. Tsang, D. S. C. Hui, M. Y. W. Kwan, W.-H. Chan, S. S. Chiu, R. L. W. Ko, K. H. Chan, S. M. S. Cheng, R. A. P. M. Perera, B. J. Cowling, L. L. M. Poon, M. Peiris, Neutralizing antibody titres in SARS-CoV-2 infections. Nat. Commun. 12, 63 (2021).

14. A. Wajnberg, F. Amanat, A. Firpo, D. R. Altman, M. J. Bailey, M. Mansour, M. McMahon, P. Meade, D. R. Mendu, K. Muellers, D. Stadlbauer, K. Stone, S. Strohmeier, V. Simon, J. Aberg, D. L. Reich, F. Krammer, C. Cordon-Cardo, Robust neutralizing antibodies to SARS-CoV-2 infection persist for months. Science. 370, 1227–1230 (2020).

15. J. M. Dan, J. Mateus, Y. Kato, K. M. Hastie, E. D. Yu, C. E. Faliti, A. Grifoni, S. I. Ramirez, S. Haupt, A. Frazier, C. Nakao, V. Rayaprolu, S. A. Rawlings, B. Peters, F. Krammer, V. Simon, E. O. Saphire, D. M. Smith, D. Weiskopf, A. Sette, S. Crotty, Immunological memory to SARS-CoV-2 assessed for up to eight months after infection. bioRxiv (2020), doi:10.1101/2020.11.15.383323.

16. A. G. L’Huillier, B. Meyer, D. O. Andrey, I. Arm-Vernez, S. Baggio, A. Didierlaurent, C. S. Eberhardt, I. Eckerle, C. Grasset-Salomon, A. Huttner, K. M. Posfay-Barbe, I. S. Royo, J. A. Pralong, N. Vuilleumier, S. Yerly, C.-A. Siegrist, L. Kaiser, Geneva Centre for Emerging Viral Diseases, Antibody persistence in the first six months following SARS-CoV-2 infection among hospital workers: a prospective longitudinal study. Clin. Microbiol. Infect. (2021), doi:10.1016/j.cmi.2021.01.005.

17. E. Duysburgh, L. Mortgat, C. Barbezange, K. Dierick, N. Fischer, L. Heyndrickx, V. Hutse, I. Thomas, S. Van Gucht, B. Vuylsteke, K. K. Ariën, I. Desombere, Persistence of IgG response to SARS-CoV-2. Lancet Infect. Dis. 21, 163–164 (2021).

18. G. den Hartog, E. R. A. Vos, L. L. van den Hoogen, M. van Boven, R. M. Schepp, G. Smits, J. van Vliet, L. Woudstra, A. J. Wijmenga-Monsuur, C. C. E. van Hagen, E. A. M. Sanders, H. E. de Melker, F. R. M. van der Klis, R. S. van Binnendijk, Persistence of antibodies to SARS-CoV-2 in relation to symptoms in a nationwide prospective study. Clin. Infect. Dis. (2021), doi:10.1093/cid/ciab172.

19. R. Verity, L. C. Okell, I. Dorigatti, P. Winskill, C. Whittaker, N. Imai, G. Cuomo-Dannenburg, H. Thompson, P. G. T. Walker, H. Fu, A. Dighe, J. T. Griffin, M. Baguelin, S. Bhatia, A. Boonyasiri, A. Cori,Z. Cucunubá, R. FitzJohn, K. Gaythorpe, W. Green, A. Hamlet, W. Hinsley, D. Laydon, G. Nedjati-Gilani, S. Riley, S. van Elsland, E. Volz, H. Wang, Y. Wang, X. Xi, C. A. Donnelly, A. C. Ghani, N. M. Ferguson, Estimates of the severity of coronavirus disease 2019: a model-based analysis. Lancet Infect. Dis. 20, 669–677 (2020).

20. E. S. Theel, P. Slev, S. Wheeler, M. R. Couturier, S. J. Wong, K. Kadkhoda, The Role of Antibody Testing for SARS-CoV-2: Is There One? J. Clin. Microbiol. 58 (2020), doi:10.1128/JCM.00797-20.

21. A. J. Jääskeläinen, S. Kuivanen, E. Kekäläinen, M. J. Ahava, R. Loginov, H. Kallio-Kokko, O. Vapalahti, H. Jarva, S. Kurkela, M. Lappalainen, Performance of six SARS-CoV-2 immunoassays in comparison with microneutralisation. J. Clin. Virol. 129, 104512 (2020).

22. P. H. Herroelen, G. A. Martens, D. De Smet, K. Swaerts, A.-S. Decavele, Humoral Immune Response to SARS-CoV-2. Am. J. Clin. Pathol. 154, 610–619 (2020).

23. A. Krüttgen, C. G. Cornelissen, M. Dreher, M. Hornef, M. Imöhl, M. Kleines, Comparison of four new commercial serologic assays for determination of SARS-CoV-2 IgG. J. Clin. Virol. 128, 104394 (2020).

24. P. Vetter, S. Cordey, M. Schibler, L. Vieux, L. Despres, F. Laubscher, D. O. Andrey, R. Martischang, S. Harbarth, C. Cuvelier, M. Bekliz, I. Eckerle, C.-A. Siegrist, A. M. Didierlaurent, C. S. Eberhardt, B. Meyer, L. Kaiser, Geneva Center for Emerging Viral Diseases, Clinical, virological and immunological features of a mild case of SARS-CoV-2 re-infection. Clin. Microbiol. Infect. (2021), doi:10.1016/j.cmi.2021.02.010.

25. J. Seow, C. Graham, B. Merrick, S. Acors, S. Pickering, K. J. A. Steel, O. Hemmings, A. O’Byrne, N. Kouphou, R. P. Galao, G. Betancor, H. D. Wilson, A. W. Signell, H. Winstone, C. Kerridge, I. Huettner, J. M. Jimenez-Guardeño, M. J. Lista, N. Temperton, L. B. Snell, K. Bisnauthsing, A. Moore, A. Green, L. Martinez, B. Stokes, J. Honey, A. Izquierdo-Barras, G. Arbane, A. Patel, M. K. I. Tan, L. O’Connell, G. O’Hara, E. MacMahon, S. Douthwaite, G. Nebbia, R. Batra, R. Martinez-Nunez, M. Shankar-Hari, J. D. Edgeworth, S. J. D. Neil, M. H. Malim, K. J. Doores, Longitudinal observation and decline of neutralizing antibody responses in the three months following SARS-CoV-2 infection in humans. Nat Microbiol. 5, 1598–1607 (2020).

26. C. Gaebler, Z. Wang, J. C. C. Lorenzi, F. Muecksch, S. Finkin, M. Tokuyama, A. Cho, M. Jankovic, D. Schaefer-Babajew, T. Y. Oliveira, M. Cipolla, C. Viant, C. O. Barnes, Y. Bram, G. Breton, T. Hägglöf, P. Mendoza, A. Hurley, M. Turroja, K. Gordon, K. G. Millard, V. Ramos, F. Schmidt, Y. Weisblum, D. Jha, M. Tankelevich, G. Martinez-Delgado, J. Yee, R. Patel, J. Dizon, C. Unson-O’Brien, I. Shimeliovich, D. F. Robbiani, Z. Zhao, A. Gazumyan, R. E. Schwartz, T. Hatziioannou, P. J. Bjorkman, S. Mehandru, P. D. Bieniasz, M. Caskey, M. C. Nussenzweig, Evolution of antibody immunity to SARS-CoV-2. Nature (2021), doi:10.1038/s41586-021-03207-w.

27. P. G. Choe, C. K. Kang, H. J. Suh, J. Jung, K.-H. Song, J. H. Bang, E. S. Kim, H. B. Kim, S. W. Park, N. J. Kim, W. B. Park, M.-D. Oh, Waning Antibody Responses in Asymptomatic and Symptomatic SARS-CoV-2 Infection. Emerg. Infect. Dis. 27 (2021), doi:10.3201/eid2701.203515.

28. W. Korte, M. Buljan, M. Rösslein, P. Wick, V. Golubov, J. Jentsch, M. Reut, K. Peier, B. Nohynek, A. Fischer, R. Stolz, M. Cettuzzi, O. Nolte, SARS-CoV-2 IgG and IgA antibody response is gender dependent; and IgG antibodies rapidly decline early on. J. Infect. 82 (2021), pp. E11–e14.

29. Hôpitaux universitaires de Genève. COVID-19 étude entreprises (SEROCoV-Work+), (2020). https://www.hug.ch/medecine-premier-recours/covid-19-etude-entreprises-serocov-work [Accessed Mar 12 2021].

30. W. J. Rogan, B. Gladen, Estimating prevalence from the results of a screening test. Am. J. Epidemiol. 107, 71–76 (1978).

31. A. Gelman, B. Carpenter, Bayesian analysis of tests with unknown specificity and sensitivity. J. R. Stat. Soc. Ser. C Appl. Stat. 69, 1269–1283 (2020).

