## Supplemental Materials for "Persistence and detection of anti-SARS-CoV-2 antibodies: immunoassay heterogeneity and implications for serosurveillance"

#### S1 Cohort description

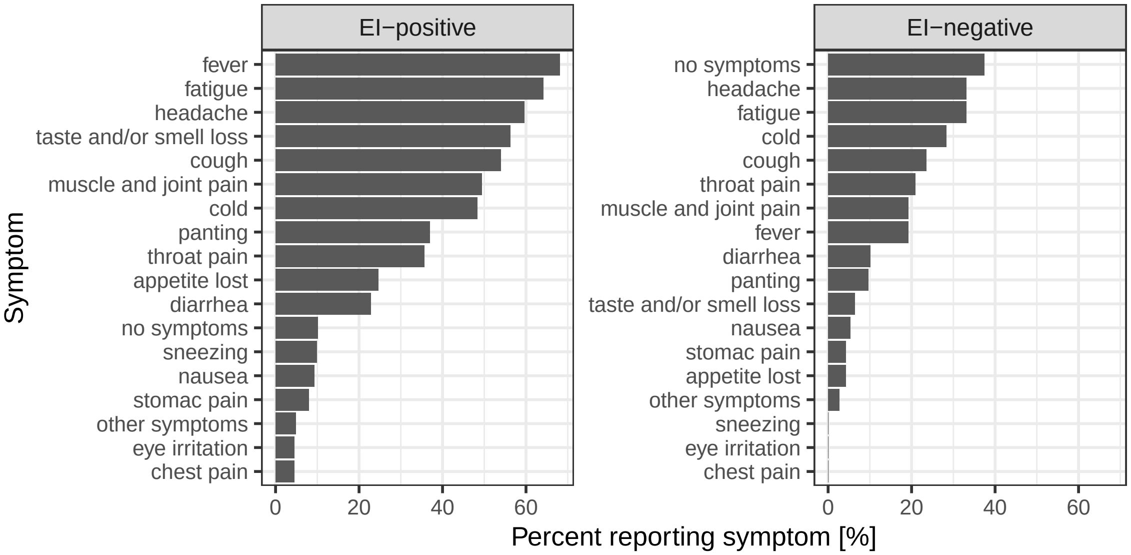


Figure S1: Self-reported symptoms before the baseline visit in each study cohort.

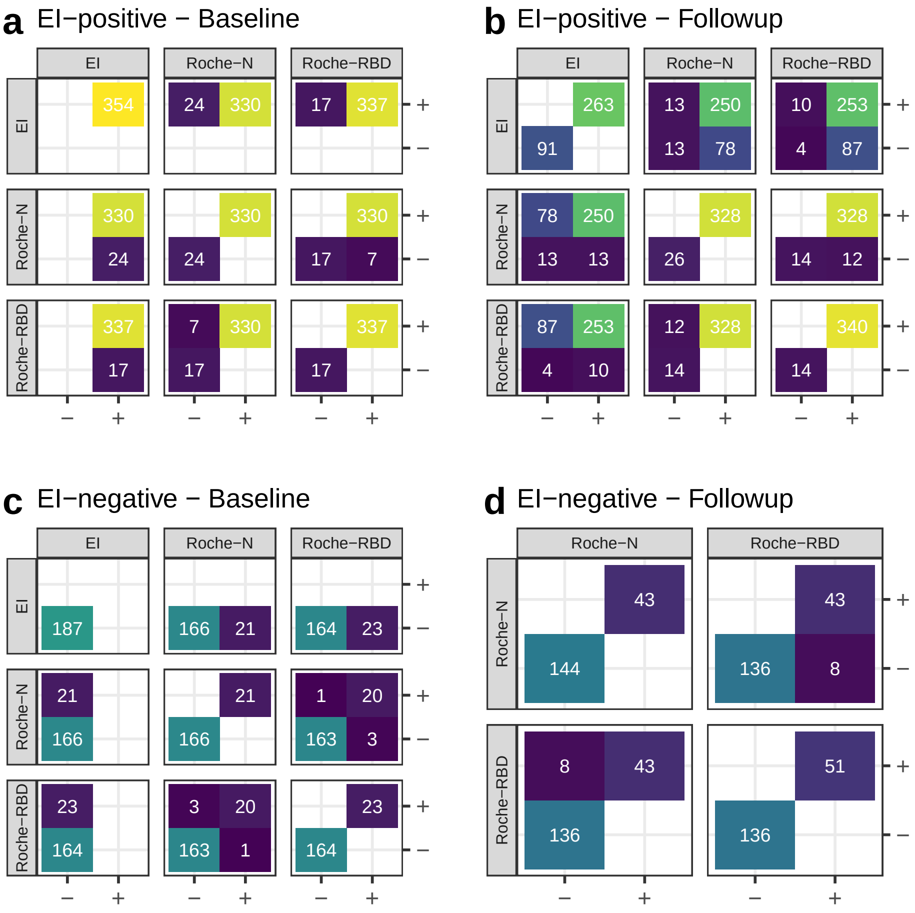


Figure S2: Two-by-two serological test confusion matrices. Results are shown for the EI-positive (a,b) and EI-negative (c,d) cohorts at baseline (a,c) and at followup (b,d). EI results were not available at followup for the negative cohort. Tests are abbreviated as EI: Euroimmun anti-S1 IgG, Roche-N: Roche anti-N total Ig, Roche-RBD: Roche anti-RBD total Ig.

Table S1: Cohort serostatus change and test response trajectories by sex and age group. Test results for the EI assay where not available for the EI-negative cohort. Significance of differences between proportions within each strata was determined by two-sided Wilson tests. Significance of trajectory increase or decrease was not computed for EI and Roche-N given that they are defined as qualitative or semi-quantitative by the manufacturers. For Roche-RBD significance was based on the z-score of baseline and followup values using a CV of 7.6%. Test abbreviations as in Figure S2.

| | Category | | EI | | Roche-N | | Roche-RBD | |
|---|---|---|---|---|---|---|---|---|
| | | | N change (%) | p-value | N change (%) | p-value | N change (%) | p-value |
| **EI-positive** | | | | | | | | |
| *Sero-status change* | | | | | | | | |
| reversion | | | | | | | | |
| | age | (17,65] | 82/313 | 0.69 | 6/293 | 0.82 | 0/298 | - |
| | | (65,105] | 9/41 | | 0/37 | | 0/39 | |
| | sex | female | 41/183 | 0.18 | 4/167 | 0.70 | 0/170 | - |
| | | male | 50/171 | | 2/163 | | 0/167 | |
| conversion | | | | | | | | |
| | age | (17,65] | - | - | 3/20 | 1.00 | 3/15 | 1.00 |
| | | (65,105] | - | | 1/4 | | 0/2 | |
| | sex | female | - | - | 3/16 | 1.00 | 2/13 | 1.00 |
| | | male | - | | 1/8 | | 1/4 | |
| *Antibody level* | | | | | | | | |
| decay | | | | | | | | |
| | age | (17,65] | - | - | - | - | 44 (14.1%) | 0.06 |
| | | (65,105] | - | | - | | 11 (26.8%) | |
| | sex | female | - | - | - | - | 15 (8.2%) | 0.0001 |
| | | male | - | | - | | 40 (23.4%) | |
| increase | | | | | | | | |
| | age | (17,65] | - | - | - | - | 199 (63.6%) | 0.17 |
| | | (65,105] | - | | - | | 21 (51.2%) | |
| | sex | female | - | - | - | - | 123 (67.2%) | 0.05 |
| | | male | - | | - | | 97 (56.7%) | |
| **EI-negative** | | | | | | | | |
| *Sero-status change* | | | | | | | | |
| reversion | | | | | | | | |
| | age | (17,65] | - | - | 3/21 | - | 1/21 | 1.00 |
| | | (65,105] | - | | 0/0 | | 0/2 | |
| | sex | female | - | - | 0/7 | 0.51 | 0/7 | 1.00 |
| | | male | - | | 3/14 | | 1/16 | |
| conversion | | | | | | | | |
| | age | (17,65] | - | - | 25/162 | 0.88 | 29/162 | 1.00 |
| | | (65,105] | - | | 0/4 | | 0/2 | |
| | sex | female | - | - | 13/86 | 1.00 | 16/86 | 0.90 |
| | | male | - | | 12/80 | | 13/78 | |
| *Antibody level* | | | | | | | | |
| decay | | | | | | | | |
| | age | (17,65] | - | - | - | - | 5 (2.7%) | 0.29 |
| | | (65,105] | - | | - | | 1 (25.0%) | |
| | sex | female | - | - | - | - | 2 (2.2%) | 0.69 |
| | | male | - | | - | | 4 (4.3%) | |
| increase | | | | | | | | |
| | age | (17,65] | - | - | - | - | 43 (23.5%) | 1.00 |
| | | (65,105] | - | | - | | 1 (25.0%) | |
| | sex | female | - | - | - | - | 21 (22.6%) | 0.90 |
| | | male | - | | - | | 23 (24.5%) | |

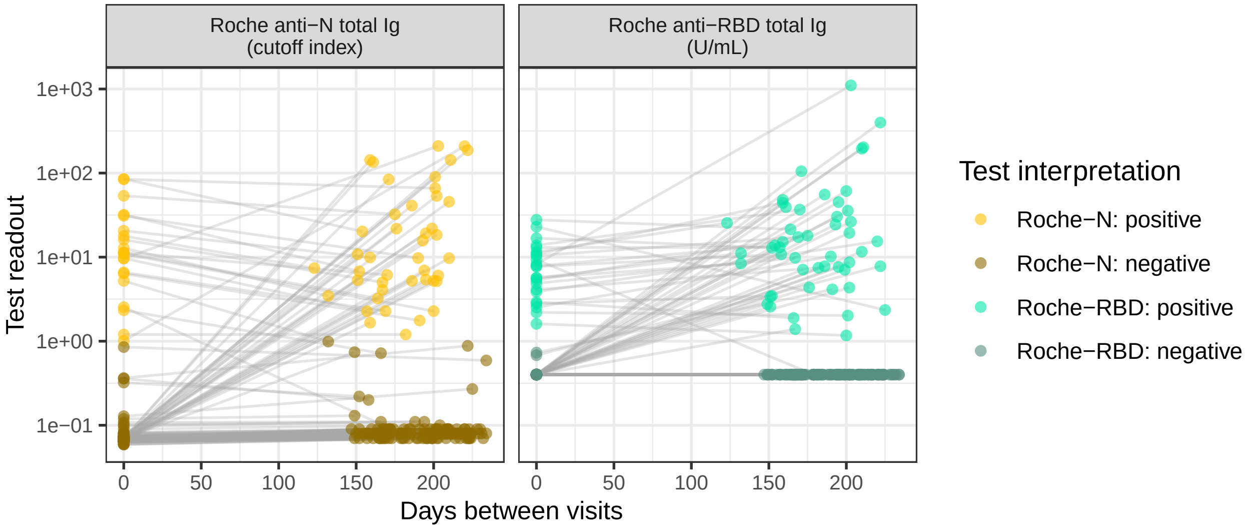


Figure S3: Test readout trajectories for the EI-negative cohort (n=187). EI results for this cohort were not available at followup. Test abbreviations as in Figure S2.

#### S2 Test readout variation

Internal quality controls for the three immunoassays (Euroimmun anti-S1, Roche anti-N and Roche anti-S) were performed using the same internal positive control, an in-house diluted leftover serum sample with high antibody levels, allowing for inter- and intra-lot comparisons for each test (determination of the coefficient of variation, CV).

All baseline and follow-up samples were tested using the same lot of Roche-N and Roche-RBD immunoassays, with intra-lot CVs of 4.66 and 7.55%, respectively. EI test readouts for our internal quality control (IQC) across lots show substantial intra- (CV range 8.06%-15.5%) as well as inter-lot variability (CV=30.4%, Fig. S4). All EI tests on follow-up samples were performed using the same lot (EI-9), which had a significantly lower IQC mean than all other lots (mean: 1.39, sd: 0.12, pair-wise p-values t-test with correction for multiple testing $< 0.01$).

We performed a sensitivity analysis on EI-based seroconvertion/reversion rates by analyzing samples processed using lot EI-7 at baseline, which had the closest IQC mean to the follow-up lot (mean: 1.66, sd: 0.22, p-value of difference with followup lot with correction for multiple testing 0.004). Baseline samples for a total of 127 participants (63 (50%) of which were female, and 111 (87%) were between 18 and 65) were processed using this lot. EI test readout trajectories and distributions at each visit are given in Fig. S5. The percentage of seroreversions in this baseline lot what 27% (31/ 115), not significantly different from the overall reversion rate in the EI-positive cohort (91/354, p-value: 0.89).

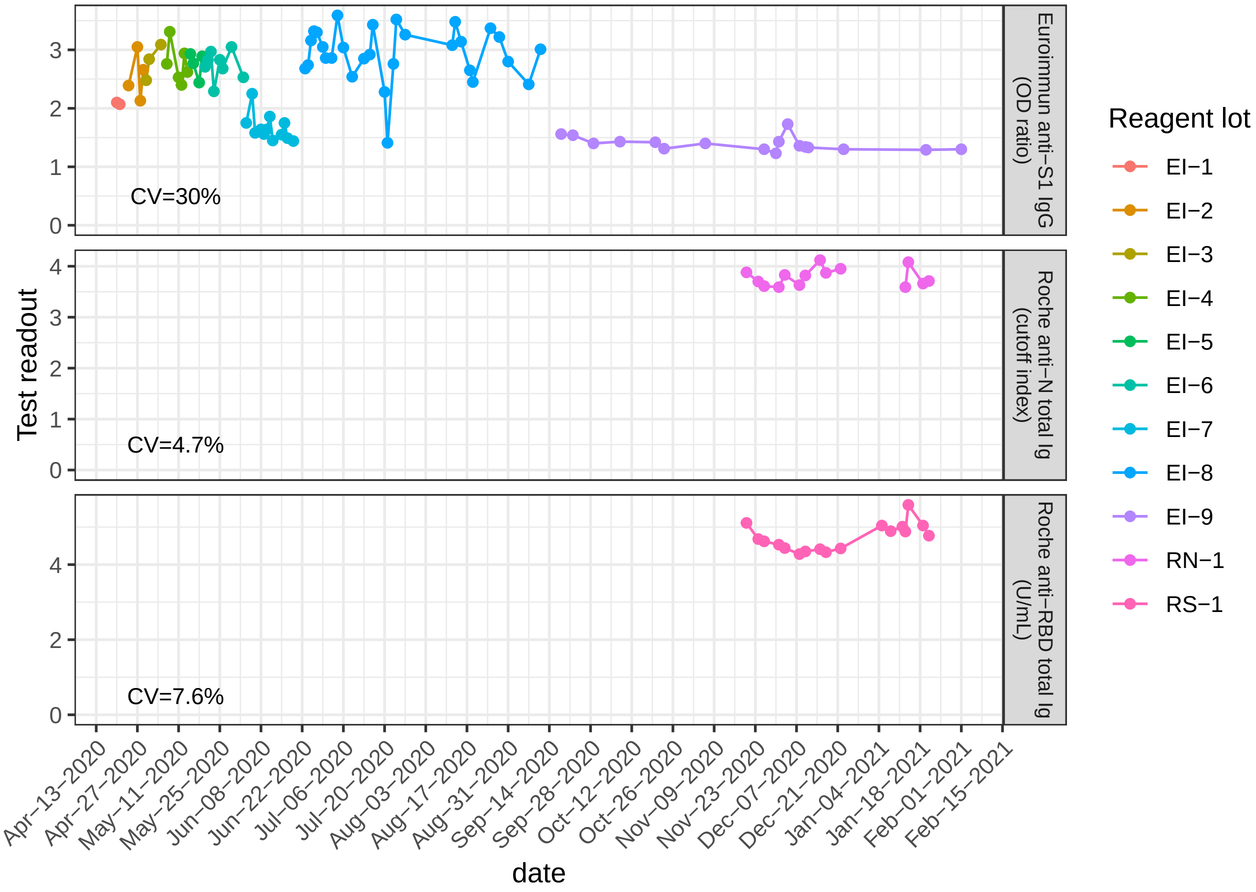


Figure S4: Test internal quality control of variation within and across test reagent lots. The IQC consisted of an in-house diluted leftover serum sample with high antibody levels. Readout units are test-specific. The inter-lot coefficient of variation (CV) is given for each test.

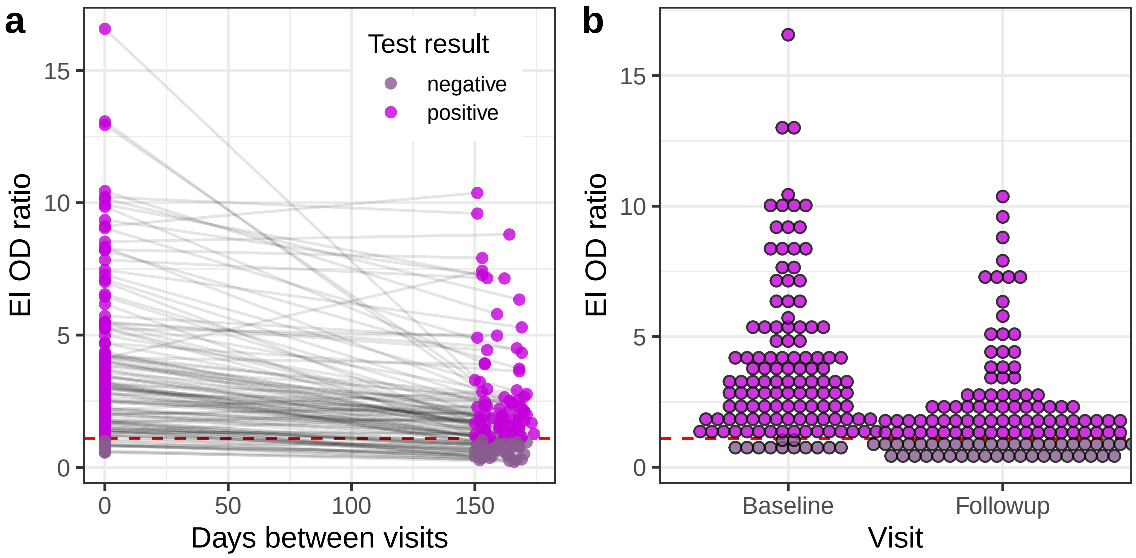


Figure S5: EI test readout trajectories and distribution in lot wih low internal control values. Changes in test readout for lot EI-7 between visits (a) and distribution for at each visit (b, dotplot with binwidth of 0.45). Horizontal dashed lines indicates the manufacturers reccomended seropositivity threshold (OD/IC ratio = 1.1).

### S3 Latent class model

Our aim is to jointly infer the underlying sero-status in both followup ('positive') and negative control cohorts and the performance (specificity and sensitivity) of multiple tests in the absence of a gold-standard for identifying historic SARS-CoV-2 infections. To do so we use a latent class model accounting for multiple imperfect tests with unknown sensitivity and specificity, and possible sero-status changes (due to infection) in between visits. Inference is made in a Bayesian framework that incorporates our longitudinal serologic data as well as in-house and external assay validation datasets.

#### S3.1 Model description

Let $z_i^0, z_i^1 \in \{0, 1\}$ be the latent class representing the true underlying infection history of participants $i$ at baseline ($z_i^0$) and at followup ($z_i^1$), with $z_i^{\cdot} = 0$ denoting infection naive participants, and $z_i^{\cdot} = 1$ those where were infected in the past. We have access to multiple observations of $z_i$ from $J$ different serological assays, $y_{i,j}, j \in \{1, \dots J\}$, both at baseline ($y_{i,j}^0$) and at followup ($y_{i,j}^1$) (Fig. S6). We assume that an individual's infection history can change in between visits due to SARS-CoV-2 infection. which occurs with probability $\mathbb{P}(\text{infection}) = \lambda$. Possible infection history trajectories are therefore; (1) remaining un-exposed from baseline to followup (with probability $1 - \lambda$),(2) going from un-exposed to infected (with probability $\lambda$), or (3) already having been infected at baseline.

#### S3.2 Time-invariant test sensitivity

Given the infection history of individual $z_i$, we model the probability of test result $y_{i,j}$ for individual $i$ and test $j$ as a Bernoulli random variable which depends on the test's specificity, $\theta^-$ (1-$\mathbb{P}$(false positive)), and sensitivity, $\theta^+$ ($\mathbb{P}$(true positive)), which we first consider to be time (since infection) invariant. We do not know the true underlying status, $z_i$. We can however obtain the probability of a serological test result given test sensitivity and specificity by marginalizing out the probability of having infection history $z_i$:

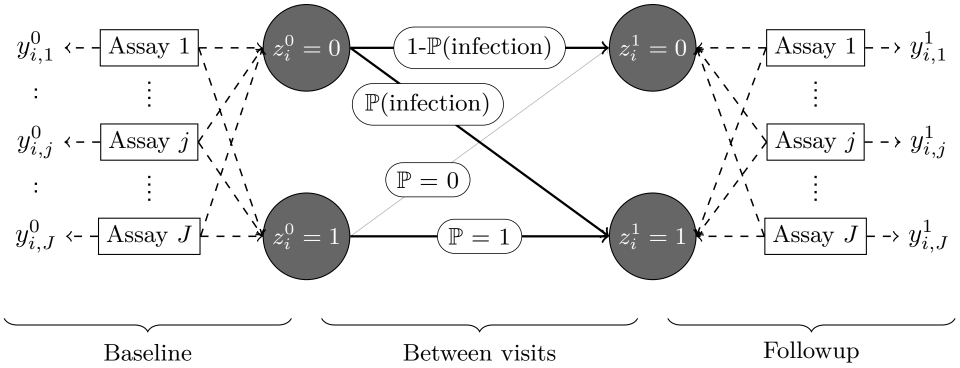


Figure S6: Latent class model diagram.

$$
\begin{aligned}
\mathbb{P}(y_{i,j}|\theta^+_j, \theta^-_j) &= \sum_{k=0,1} \mathbb{P}(y_{i,j}|z_i = k, \theta^+_j, \theta^-_j) \times \mathbb{P}(z_i = k) \\
&= \sum_{k=0,1} Bernoulli(y_{i,j}|z_i = k, \theta^+_j, \theta^-_j) \times \mathbb{P}(z_i = k) \\
&= \sum_{k=0,1} (k\theta^+_j + (1-k)(1-\theta^-_j))^{y_{i,j}} (1 - k\theta^+_j + (1-k)(1-\theta^-_j))^{1-y_{i,j}} \times \mathbb{P}(z_i = k).
\end{aligned}
$$

Each individual has an unknown underlying infection status at baseline ($z^0_i$), which can change at followup ($z^1_i$) due to infection in between visits (Fig.S6). Assuming test sensitivity and specificity do not vary with time post-infection, the probability of a baseline-followup pair of observations $\{y^0_{i,j}, y^1_{i,j}\}$ can be obtained by marginalizing out both infection-statuses at baseline and followup as:

$$
\begin{aligned}
\mathbb{P}(y^0_{i,j}, y^1_{i,j}|\theta^+_j, \theta^-_j) = \sum_{k=0,1}\sum_{l=0,1} &\mathbb{P}(y^0_{i,j}|z^0_i = k, \theta^+_j, \theta^-_j) \times \mathbb{P}(z^0_i = k) + \\
&\mathbb{P}(y^1_{i,j}|z^1_i = l, \theta^+_j, \theta^-_j) \times \mathbb{P}(z^1_i = l|z^0_i = k)\mathbb{P}(z^0_i = k),
\end{aligned}
\tag{1}
$$

where $\mathbb{P}(z^1_i = l|z^0_i = k)$ is the conditional probability of having infection history $l$ at followup status $k$ at baseline. Following the description in Fig.S6, we have four possible combinations of baseline-followup infection histories with probabilities:

$$
\mathbb{P}(z^1_i = l|z^0_i = k)\mathbb{P}(z^0_i = k) = \begin{cases} (1-\lambda)(1-\rho) & \text{if } k = 0 \text{ and } l = 0 \\ \lambda(1-\rho) & \text{if } k = 0 \text{ and } l = 0 \\ \rho & \text{if } k = 1 \text{ and } l = 1 \\ 0 & \text{if } k = 1 \text{ and } l = 0 \end{cases},
$$

where $\mathbb{P}(z^0_i = 1) = \rho$ is the prior probability of individual $i$ being sero-positive at baseline.

For a set of $J$ different serological tests, the likelihood of parameter set $\boldsymbol{\Theta} = \{\boldsymbol{\theta^+}, \boldsymbol{\theta^-}, \boldsymbol{\eta}, \rho, \lambda\}$, where $\boldsymbol{\theta}^{+(-)} = \{\theta_1^{+(-)}, \ldots, \theta_J^{+(-)}\}$ is the vector of all test sensitivities (specificities), given observations $\{\boldsymbol{y^0}, \boldsymbol{y^1}\} = \{y^0_{1,\ldots,i,\ldots,N,1\ldots,j,\ldots J}, y^1_{1,\ldots,i,\ldots,N,1\ldots,j,\ldots J}\}$ is:

$$
\mathcal{L}(\boldsymbol{\Theta}|\boldsymbol{y^0}, \boldsymbol{y^1}) = \prod_{i=1}^{N}\prod_{j=1}^{J} \mathbb{P}(y^0_{i,j}, y^1_{i,j}|\theta^+_j, \theta^-_j). \tag{2}
$$

#### S3.3 Time-varying test sensitivity

In eq.1 we assume that test sensitivity and specificity are the same at baseline and followup. While by definition, specificity can not vary with time since infection, we expect that test sensitivity will decrease as the time from infection increases (after the acute-convalescent period) due to the decay of circulating antibodies.

Instead of having a single value of sensitivity for serological test $j$, we can make it depend on time post infection (tpi) $\tau$, $\theta^+_{j,\tau}$. Since the tpi is not usually available we therefore need to marginalize over possible delays between infection and serological visits:

$$\mathbb{P}(y_{i,j}|\theta^+_{j,\tau}, \theta^-_j) = \sum_{\tau_i=0}^{\tau_i^{max}} \sum_{k=0,1} Bernoulli(y_{i,j}|z_i = k, \theta^+_{j,\tau_i}, \theta^-_j) \times \mathbb{P}(z_i = k) \times \mathbb{P}(\tau_i),$$

where $\mathbb{P}(\tau_i)$ is the probability of participant $i$ having been infected $\tau_i$ days prior to the visit date, $t_i^v$, and $\tau_i^{max}$ is the longest possible delay given the start of the pandemic at $t_0$.

Following (Azman et al. 2020), we model time-varying sensitivity as a cubic polynomial of $\log(\tau)$ on the logit-scale:

$$\text{logit}(\theta^+_{j,\tau}) = \alpha_j + \beta_{1,j}\log(\tau) + \beta_{2,j}\log(\tau)^2 + \beta_{3,j}\log(\tau)^3. \quad (3)$$

To set $\mathbb{P}(\tau_i)$ we assume that the probability of infection on a given day $t_i^s$ is proportional to the daily number of reported virologically-confirmed SARS-CoV-2 infections (in Geneva) accounting for the delay between infection, symptom onset, and case reporting, $\delta$:

$$\mathbb{P}(\tau_i) = \mathbb{P}(t_i^s = t|t_i^v) = \frac{cases_{t+\delta}}{\sum_{t\prime=t_0}^{t_i^v} cases_{t'+\delta}}.$$

Time between infection and symptom onset was based on median estimates of the incubation period and set to 5 days (Lauer et al. 2020). The delay between cases and symptom onset was set to 6 days for the first wave (Sciré et al. 2020), which we assumed halved during the second pandemic wave due to test capacity buildup (Stringhini, Zaballa, et al. 2021). Given strong intra-week case reporting variability we take a 7-day moving average of reported cases to compute $\mathbb{P}(\tau_i)$.

#### S3.4 Test performance validation datasets

Following our previous seroprevalence estimation framework in (Stringhini, Wisniak, et al. 2020), we treat test sensitivity and specificity as unknown and use validation data to inform their values (Gelman and Carpenter 2020). To do so we use both test-specific validation datasets, as well as a validation dataset produced by the virology laboratory at Geneva University Hospitals (HUG) for which all three immunoassays were run on each sample.

##### S3.4.1 Test-specific validation data

For test-specific validation study $s$, test specificity is informed by the number of false positives, $n^-_{s,j}$, resulting from a negative control sample of size $N^-_{s,j}$, modeled as a binomial distribution:

$$n^-_{s,j} \sim Binomial(N^-_{s,j}, 1 - \theta^-_{s,j}).$$

Numbers and data sources for test-specific validation datasets are given in Table S2.

For sensitivity validation studies we exploit available information on the likely time post infection to inform time-varying sensitivity. While exact infection dates are typically not known (or reported), data on the number of true positives within a range $k$ of times post symptom onset or post RT-PCR test, $\boldsymbol{\tau}'_k = \{\tau'_{min,k}, \ldots, \tau'_{max,k}\}$, $n^+_{s,j,[\tau_{min}-\tau_{max}]}$, among $N^-_{s,j,\boldsymbol{\tau}'_k}$ positive controls:

$$\mathbb{P}(n^+_{s,j,\boldsymbol{\tau}'_k} | N^+_{s,j,\boldsymbol{\tau}'_k}, \alpha_{j,s}, \beta_{j,1-3}) = \frac{1}{\tau^k_{max} - \tau^k_{min}} \sum_{\tau=\tau^k_{min}}^{\tau^k_{max}} Binomial(N^+_{s,j,\boldsymbol{\tau}'_k}, \theta^+_{s,j,\tau}),$$

where time varying-sensitivity $\theta^+_{s,j,\tau}$ is given as in eq.3, with the intercept $\alpha_{j,s}$ allowed to vary between studies (as opposed to polynomial coefficients $\beta_{1-3}$ that are assumed to be the same across studies for a given test). When validation studies report times post symptom onset we extend the ranges by the median incubation period of SARS-CoV-2, $\delta_{inc} = 5$ days, (Lauer et al. 2020) ($\tau_{min/max} = \tau'_{min/max} + \delta_{inc}$). When delays are expressed as times post RT-PCR test we assume that symptom onset may have occurred up to 10 days prior to testing, and thus extend the upper bound of the range by that amount ($\tau_{max} = \tau'_{max} + \delta_{inc} + 10$). We assume that there is equal probability of delays within each range $\boldsymbol{\tau}'_k$. Time-varying sensitivity validation data used in the analysis are shown in Fig. S7.

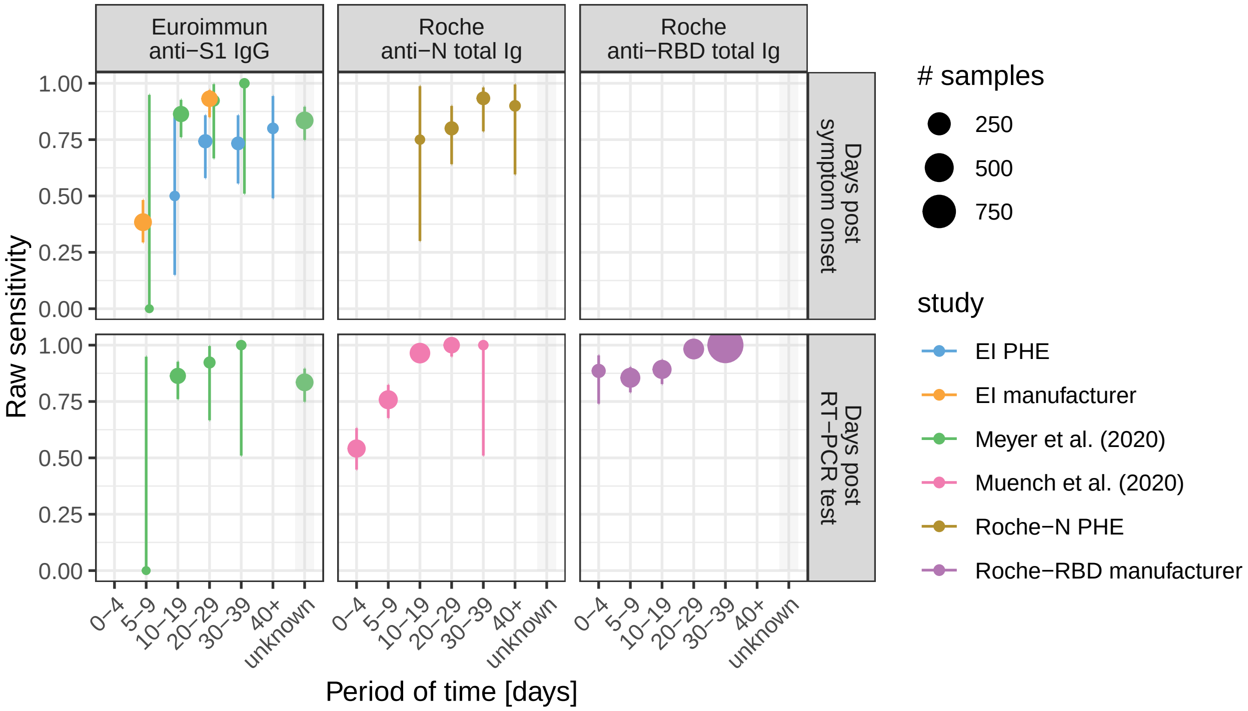


Figure S7: Time-informed sensitivity validation studies. Raw sensitivity (percen positive among positive controls) are given with Wilson binomial 95% confidence intervals. and Study sources given in Table S2, in addition to evaluation reports from Public Health England on EI (PHE 2020b) (also contains manufacturer data), and Roche-N (PHE 2020a).

##### S3.4.2 Multi-assay validation data

Validation data covering all three test in each participant provide information on joint test performance. Assuming test results are independent from each other, the probability of a set of results $\{y^c_{i,1}, \ldots, y^c_{i,J}\}$ for control participant $i$ with known infection history $z^c_i$ is given by:

Table S2: Test-specific validation studies

| | **Specificity** | | **Sensitiviy** | | |
|---|---|---|---|---|---|
| Test | N controls | N false positives | N controls | N true positives | source |
| Euroimmun anti-RBD IgG | 326 | 4 | 181 | 154 | Meyer et al. 2020 |
| Roche anti-N total Ig | 10453 | 21 | 185 | 184 | Muench et al. 2020 |
| Roche anti-RBD total Ig | 5991 | 1 | 1423 | 1406 | Roche 2020 |

$$\begin{aligned}\mathbb{P}(y^c_{i,1},\ldots,y^c_{i,J}|\boldsymbol{\theta^+}_\tau,\boldsymbol{\theta^-},z_i=z^c_i,\tau) &= \prod_{j=1}^{J}\mathbb{P}(y^c_{i,j}|\theta^+_{j,\tau},\theta^-_j,z^c_i)\\ &= \prod_{j=1}^{J} Bernoulli(y^c_{i,j}|\theta^+_{j,\tau},\theta^-_j,z_i=z^c_i).\end{aligned}$$

#### S3.5 Hierarchical Bayesian inference framework

We combine our longitudinal data, external test-specific validation data and the in-house multi-assay validation dataset in a hierarchical Bayesian framework that allows for pooling between separate sensitivity and specificity estimates (Gelman and Carpenter 2020).

Pooling between external case-specific and the multi-assay validation studies was implemented by modeling specificity and sensitivity of each test $j$ and study $s$ on the logit scale:

$$\begin{aligned}\text{logit}(\alpha_{j,s}) &\sim \mathcal{N}(\mu_{\alpha_j},\sigma_{\alpha_j})\\ \text{logit}(\theta^-_{j,s}) &\sim \mathcal{N}(\mu_{\theta^-_j},\sigma_{\theta^-_j})\\ \mu_{\theta^-_j} &\sim \mathcal{N}(4,2)\\ \mu_{\alpha_j} &\sim \mathcal{N}(2,1)\\ \sigma_{\theta^-_j} &\sim \mathcal{N}^+(0,1)\\ \sigma_{\alpha_j} &\sim \mathcal{N}^+(0,1),\end{aligned}$$

where $\mu_{\theta^-_j}$ and $\sigma_{\theta^-_j}$ are hyperparameters corresponding to the logit-scale pooled mean and variance of the specificity of test $j$, and $\mathcal{N}^+$ is the positive half-normal distribution. Similarly we pool the estimates of the intercept of the cubic polynomial modeling sensitivity (eq. 3), with test-specific mean $\mu_{\alpha_j}$ and standard deviation $\sigma_{\alpha_j}$. Following (Gelman and Carpenter 2020) we choose weekly informative priors for $\sigma_{\theta^-},\sigma_\alpha$ which allows for weak pooling while letting significant variation between study estimates, and for $\mu_{\theta^-}$ and $\mu_\alpha$ which put two-thirds of the mass in the intervals $(0.881, 0.997)$ and $(0.731, 0.952)$ respectively.

The latent class model also allows for the inference of the probability of infection between visits, $\lambda$, for which we give un-informative priors between 0 and 1. We also set weakly informative normal priors on the time-varying sensitivity polynomial coefficients $\beta_{1-3,j} \sim \mathcal{N}(0,1)$, and a strong prior on the sensitivity at 0 days post infection $\theta^+_{0,j} \sim \mathcal{N}(0, 0.01)$ representing the fact that there is a lag between infection and immune response buildup against SARS-CoV-2.

The prior probability of being infected, $\rho$, is set separately for each participants using information on eventual SARS-CoV-2 test results during the study period. We assume the prior of being infected at baseline of a participant had an RT-PCR positive result prior to the baseline visit was equal to the test specificity, here assumed to be 98%. Conversely, if the participant reported having an RT-PCR positive result between the first and second visit, then the prior probability of being infected at baseline was of 2% (= 1− test specificity). The prior was set to 50% if no diagnostic test data was available.

Posterior draws were obtained using a Hamiltonian Monte Carlo sampler as implemented in the Stan programming language (Carpenter et al. 2017), through the package `rstan` (Stan Development Team 2020) in R. We ran four chains in parallel with 250 warmup iterations and 1000 sampling iterations each. Chain convergence was assessed using the Gelman-Rubin $\hat{R}$ statistic (Gelman, Rubin, et al. 1992). The code used in the analysis is available at https://github.com/UEP-HUG/serosuivi-public.

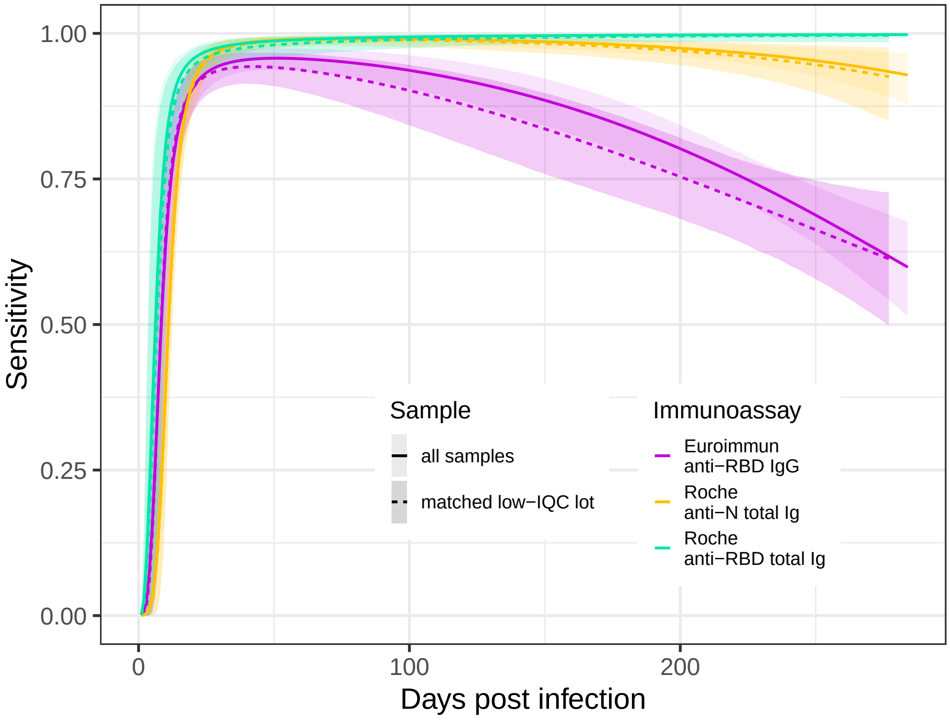


Figure S8: Time-varying estimates of test sensitivity for the whole sample and the low-IQC lot.

#### S4 Simulation analysis

To quantify the impact of time-varying sensitivity on seroprevalence estimates when using conventional methods for test performance correction. For each simulation seroprevalence level (varying between 10% and 90%) and epidemic scenario (single wave with early and late serosurvey, and two waves) described in the Materials and Methods section of the main text, we generate a set of 2000 test results based on the estimates of test specificity and time-varying sensitivity resulting from the analysis descried above.

We then estimate seroprevalence following a Bayesian hierarchical framework that allows for uncertain test performance as described in Gelman and Carpenter 2020, which we define as the "conventional" approach. Given the focus on changes in test sensitivity with time post infection, we treat specificity as a known and fix its value to the one used to generate the simulated test results. Test sensitivity, $\theta_j^+$, is treated as unknown and time-invariant, for which we pool estimates from other available studies, $s$, as detailed in Table S2, using the same priors as described in section S3.5. Given simulated test results $y_{i,j}$, seroprevalence is $p$ then estimated as:

$$
\begin{aligned}
y_{i,j} &\sim Binomial(p\theta_j^+ + (1-p)(1-\theta_j^-)) \\
\text{logit}(\theta_{j,s}^+) &\sim \mathcal{N}(\mu_{\theta_j^+}, \sigma_{\theta_j^+}) \\
\mu_{\theta_j^+} &\sim \mathcal{N}(4,2) \\
\sigma_{\theta_j^+} &\sim \mathcal{N}^+(0,1).
\end{aligned}
$$

Parameter posterior samples were drawn using the same approach as in section S3.5.

### S5 Specchio-COVID19 study group

Isabelle Arm-Vernez, Andrew S Azman, Fatim Ba, Delphine Bachmann, Jean-François Balavoine, Michael Balavoine, Lorenzo Barbieri, Jonathan Barbolini, Gil Barbosa Monteiro, Hélène Baysson, Lison Beigbeder, Sarah Emilie Berthoud, Patrick Bleich, Livia Boehm, Viola Bucolli, François Chappuis, Prune Collombet, Delphine Courvoisier, Alain Cudet, Laura Isabel Da Silva, Léah Davister, Elodie de Jesus, Carlos de Mestral Vargas, Steven Del Riccio, Paola D'ippolito, Celine Dubas, Richard Dubos, Anik Dubost, Roxane Dumont, Isabella Eckerle, Nacira El Merjani, Antoine Flahault, Natalie Francioli, Marion Frangville, Miguel Frias, Fanny Golaz, Carole Grasset Salomon, Idris Guessous, Séverine Harnal, Laurent Kaiser, Omar Kherad, Manon Ladouce, Catarina Ines Leite Alves, Pierre Lescuyer, François L'Huissier, Andrea Jutta Loizeau, Fanny-Blanche Lombard, Elsa Lorthe, Line Ange Mégane Maiveke, Yasmina Malim, Chantal Martinez, Lucie Ménard, Lakshmi Menon, Ludovic Metral-Boffod, Benjamin Meyer, Alexandre Moulin, Mayssam Nehme, Natacha Noël, Princess Ojo Osarugue, Valérie Pascoli, Francesco Pennacchio, Javier Perez-Saez, Attilio Picazio, Giovanni Piumatti, Didier Pittet, Jane Portier, Klara M Posfay-Barbe, Géraldine Poulain, Caroline Pugin, Nick Pullen, Zo Francia Randrianandrasana, Adrien Jos Rastello, Aude Richard, Viviane Richard, Frederic Rinaldi, Irine Sakvarelidze, Lilas Salzmann-Bellard, Kadija Samir, Silvia Stringhini, Stéphanie Testini, Camille Tible, Didier Trono, Charlotte Verolet, Guillemette Violot, Nicolas Vuilleumier, Loic Widmer, Manon Will, Ania Wisniak, Sabine Yerly, María-Eugenia Zaballa

### References


Azman, Andrew S et al. (2020). "Vibrio cholerae O1 transmission in Bangladesh: insights from a nationally representative serosurvey". In: *The Lancet Microbe* 1.8, e336–e343.

Carpenter, Bob et al. (2017). "Stan: A probabilistic programming language". In: *Journal of statistical software* 76.1.

Gelman, Andrew and Bob Carpenter (2020). "Bayesian analysis of tests with unknown specificity and sensitivity". In: *Journal of the Royal Statistical Society: Series C (Applied Statistics)* 69.5, pp. 1269–1283.

Gelman, Andrew, Donald B Rubin, et al. (1992). "Inference from iterative simulation using multiple sequences". In: *Statistical science* 7.4, pp. 457–472.

Lauer, Stephen A et al. (2020). "The incubation period of coronavirus disease 2019 (COVID-19) from publicly reported confirmed cases: estimation and application". In: *Annals of internal medicine* 172.9, pp. 577–582.

Meyer, Benjamin et al. (2020). "Validation of a commercially available SARS-CoV-2 serological immunoassay". In: *Clinical microbiology and infection* 26.10, pp. 1386–1394.

Muench, Peter et al. (2020). "Development and validation of the Elecsys Anti-SARS-CoV-2 Immunoassay as a highly specific tool for determining past exposure to SARS-CoV-2". In: *Journal of Clinical Microbiology* 58.10.

PHE (June 2020a). *Evaluation of Roche Elecsys Anti-SARS-CoV-2 serology assay for the detection of anti-SARS-CoV-2 antibodies.* Public Health England. Wellington House 133-155 Waterloo Road, London SE1 8UG, United Kindom. URL: https://assets.publishing.service.gov.uk/government/uploads/system/uploads/attachment_data/file/891598/Evaluation_of_Roche_Elecsys_anti_SARS_CoV_2_PHE_200610_v8.1_FINAL.pdf.

– (June 2020b). *Evaluation of the Euroimmun Anti-SARS-CoV-2 ELISA (IgG) serology assay for the detection of anti-SARS-CoV-2 antibodies.* Public Health England. Wellington House 133-155 Waterloo Road, London SE1 8UG, United Kindom. URL: https://assets.publishing.service.gov.uk/government/uploads/system/uploads/attachment_data/file/893433/Evaluation_of_Euroimmun_SARS_CoV_2_ELISA_IgG__1_.pdf.

Roche (2020). *Elecsys® Anti-SARS-CoV-2 S. Immunoassay for the quantitative determinationof antibodies to the SARS-CoV-2 spike protein.* Package Insert 2020-12, V1.0; Material Numbers 09289267190 and 09289275190. Roche Diagnostics International Ltd. CH-6343 Rotkreuz, Switzerland. URL: https://diagnostics.roche.com/content/dam/diagnostics/Blueprint/en/pdf/cps/Elecsys-Anti-SARS-CoV-2-S-factsheet-SEPT-2020-2.pdf.

Sciré, Jérémie et al. (2020). "Reproductive number of the COVID-19 epidemic in Switzerland with a focus on the Cantons of Basel-Stadt and Basel-Landschaft". In: *Swiss Medical Weekly* 150.19-20, w20271.

Stan Development Team (2020). *RStan: the R interface to Stan.* R package version 2.19.3. URL: http://mc-stan.org/.

Stringhini, Silvia, Ania Wisniak, et al. (2020). "Seroprevalence of anti-SARS-CoV-2 IgG antibodies in Geneva, Switzerland (SEROCoV-POP): a population-based study". In: *The Lancet* 396.10247, pp. 313–319.

Stringhini, Silvia, Maria-Eugenia Zaballa, et al. (2021). "Seroprevalence of anti-SARS-CoV-2 antibodies after the second pandemic peak". In: *The Lancet Infectious Diseases.*
